# Heart failure with physical frailty is associated with inflammation, insulin resistance, GDF-15 and impaired energy and amino acid metabolism

**DOI:** 10.1101/2025.08.26.25334502

**Authors:** Konstantinos Prokopidis, Sima Jalali Farahani, Beyza Gulsah Altinpinar, Omid Khaiyat, Adam Burke, Amy Nortcliffe, Gregory Y.H. Lip, Rajiv Sankaranarayanan, Howbeer Muhamadali, Masoud Isanejad

## Abstract

**Aims:** While physical frailty is a common feature in heart failure (HF), less is known about how frailty is linked to impaired clinical biomarkers and metabolic dysregulation in HF.

**Methods:** We recruited 25 patients with HF (67.9 ± 10.0 years) and 29 adults without HF (NonHF) (67.8 ± 11.1 years). Physical frailty was assessed using low physical activity levels with low handgrip strength (HGS) and/or 30-second chair stand test (30CST). Untargeted plasma metabolomic profiling was performed via gas chromatography-mass spectrometry. Statistical analyses were conducted via SPSS and MetaboAnalyst.

**Results:** HF-Frail (n=25) compared to NonHF-NonFrail (n=18), had lower 6-minute walking distance (386.4 vs. 501.3 meters, P<0.01) and weaker HGS/body mass index (BMI) (1.05 vs. 1.41, P=0.04). HF-Frail compared to NonHF-NonFrail had significantly elevated plasma N-terminal fragment of pro-brain natriuretic peptide (NT-proBNP) (241.2 vs. 117.1 pg/ml, P=0.007) and growth differentiation factor-15 (GDF-15) levels (1227.2 vs. 382.5 pg/ml, P<0.01). Agnostic principal component analysis revealed elevated plasma branched chain amino acids and reduced glutamine, methionine and tryptophan in HF-Frail vs. NonHF-NonFrail controls (P<0.05). Compared to HF-NonFrail (n=7), HF-Frail had lower galacturonic acid-1-phosphate, methionine, indole-3-acetamide, pyruvic and malic acid (P<0.05). Significant negative correlations were found between NT-proBNP, tumour necrosis factor-alpha (TNF-α), GDF-15, and frailty outcomes (HGS/BMI, 30CST; P<0.05).

**Conclusions:** In HF, physical frailty is linked to impaired energy and amino acid metabolism, along with elevated inflammation and GDF-15. These findings warrant for longitudinal studies to unravel clinical biomarkers that could serve as therapeutic agents, targeting frailty progression in HF.

**Lay Summary:** This study investigated the association between physical frailty, clinical biomarkers, and metabolic dysregulation in patients with heart failure (HF), revealing significant impairments in physical performance and metabolic profiles compared to non-frail HF and non-HF controls.

- Patients with HF and frailty exhibited reduced 6-minute walk distance and 30-second chair stand repetitions, had weaker handgrip strength, and elevated levels of N-terminal pro-B-type natriuretic peptide and growth differentiation factor-15 compared to non-frail controls (P<0.05).
- Metabolomic analysis showed increased plasma branched-chain amino acids, reduced glutamine and methionine, and altered energy metabolism intermediates (i.e., pyruvic acid) (P<0.05) in HF with frailty, indicating significant metabolic disruptions.

## Introduction

Heart failure (HF) is a progressive clinical cardiometabolic syndrome, imposing a significant burden on healthcare systems, particularly among ageing populations (Shahim et al., 2023). Muscle weakness, physical frailty and sarcopenia are increasingly recognized as highly common non-cardiac comorbidities in HF(Ge et al., 2025). Physical frailty, characterized by impaired muscle health (reduced strength and endurance), affects both HF with reduced ejection fraction (HFrEF) and HF with preserved ejection fraction (HFpEF), and significantly reduces the quality of life, exercise tolerance, and increasing mortality risk (Von Haehling et al., 2017). In a meta-analysis, we showed that presence of low muscle strength was linked to increased severity of HF, as indicated by higher levels of B-type natriuretic peptide (BNP) and its N-terminal fragment (NT-proBNP) (Prokopidis et al., 2024). In addition, impaired muscle health in HF may be due to systemic metabolic dysfunction rather than ageing alone (Zamani et al., 2021), but few studies have addressed these specific mechanisms.

Metabolomics coupled with biochemical blood biomarkers are promising tool to explore the molecular underpinnings of diseased states, including HF (Hahn et al., 2024) (Fuller et al., 2023). Previous studies employing metabolomics, has shown alterations in energy, amino acid, and lipid metabolism in HF, reflecting the systemic metabolic perturbations driven by cardiac dysfunction (Hage et al., 2020). However, knowledge on clinical factor contributing to these metabolic dysregulations are less known. Previously, metabolomic studies have investigated the differences of metabolic profile in HF-with diabetes and HF-with obesity, yielding important observations on the biology of diseases. Recently, there has been increasing emphasis on a comprehensive approach that integrates plasma biomarkers, metabolomic profiles, and clinical data to better understand and disentangle the common mechanisms underlying physical frailty in HF (Hahn et al., 2024).

In this context, clinical biomarkers including transforming growth factor (TGF)-β, and a group of pleiotropic cytokines, particularly activins and growth differentiation factors (GDF), such as GDF-15 and GDF-8 (or myostatin) have been implicated in various pathological conditions including cardiovascular disease(Wang et al., 2021) and muscle weakness (Kamper et al., 2024). In addition, factors such as follistatin-3 antagonists that bind activins/myostatin and prevent them from receptor signalling, may possess a key role in energy metabolism(Bielka et al., 2023). In this study, we provide novel insight into link between muscle weakness, physical frailty with important clinical bookmakers, couple plasma metabolomics to provide further insight into marked metabolism abnormalities in HF with and without physical frailty.

## Methods

### Study population

Patients with chronic HF were recruited by reviewing the medical records from Aintree Hospital at the Liverpool Hospital Foundation Trust from November 2023 until August 2024. All patients with HF have been diagnosed by cardiologists according to European Society Cardiology for HF guidelines (McDonagh et al., 2021). The NT-proBNP threshold was adapted from the National Institute for Health and Care Excellence, and echo ejection fraction estimation using the European Society Cardiology HF guidelines.

Participants had a clinical diagnosis of HF using left ventricular ejection fraction (LVEF%) for classification. Patients were included if they: i) had a body mass index (BMI) between 18 and 30 kg/m², ii) were age >50 years old, iii) were on optimal medical therapy for at least three months before study inclusion, iv) were able to provide consent and walk a minimum of 10 meters, with or without a walking aid, while v) NonHF adults were eligible if they were under control for hypertension or hypercholesterolemia without signs or history of a chronic conditions and were community-dwelling.

Participants were excluded if they: i) had a recent treatment (within three months) of antibiotics or probiotics, ii) had comorbidities such as chronic irritable bowel syndrome, kidney failure, or cancer at late stages, iii) were being treated with immunosuppressive medications, iv) had a presence of concurrent infection, and v) received cardiac resynchronization therapy within the past six months. NonHF participants were recruited from the general healthy population, self-reported no chorionic diseases, Liverpool, UK. This study received sponsorship from the University of Liverpool and ethical approval from the London-Queen Square Research Ethics Committee (REC Reference: 23/PR/0050). The study was registered at Clinicaltrials.gov UoL001725. All participants provided written informed consent at enrolment.

### Assessment of body composition, muscle strength, and physical function

The descriptive protocol is available in **Supplementary Materials**. Dual x-ray absorptiometry (GE Lunar Prodigy) was used to assess body composition. Appendicular lean soft tissue (ALST) was calculated by summing lean mass from arms and legs, excluding fat and bone, representing skeletal muscle mass.

Muscle strength was measured through handgrip strength (HGS) using a hand dynamometer (JAMAR hydraulic dynamometer), while knee extension strength was assessed by knee extension (Dynamo by VALD). The highest value of maximum three attempts was selected from the participants’ dominant arm/leg. Physical function was assessed via 10-m walking speed, 3-m timed-up-and-go testing (TUG), 6-minute walking distance (6MWD), and 30-second chair stand test (30CST).

### Physical activity assessment

Self-reported physical activity levels were assessed using the International Physical Activity Questionnaire (IPAQ) Short Form, and scores were calculated in accordance with the IPAQ scoring protocol (2005)(Hagströmer et al., 2006). Physical activity was quantified both continuously and categorically. For the continuous score, activity is expressed in metabolic equivalent (MET)-minutes per week. This is calculated by multiplying the number of minutes per day by the number of days per week and the assigned MET value for each activity domain: 3.3 for walking, 4.0 for moderate activity, and 8.0 for vigorous activity. The total physical activity score is obtained by summing all MET-minutes across the domains. For the categorical score, participants were classified into one of three levels of physical activity. Low: No reported activity, or activity levels insufficient to meet the criteria for the moderate or high categories. Moderate: Vigorous activity on ≥3 days for ≥20 minutes/day, or Moderate activity and/or walking on ≥5 days for ≥30 minutes/day, or Any combination of walking, moderate, or vigorous activity on≥5 days achieving at least 600 MET-minutes/week. High: Vigorous activity on ≥3 days accumulating at least 1,500 MET-minutes/week, or a combination of activities on ≥7 days achieving at least 3,000 MET-minutes/week.

### Physical frailty assessment

To ascertain physical frailty, we have adapted Fried criteria of frailty(Fried et al., 2001), for the low physical activity (IPAQ<2) as a component for physical frailty. Further, low HGS (men<27 kg as per the European Working Group on Sarcopenia in Older People (EWGSOP2) criteria and women<16 kg) (Cruz-Jentoft et al., 2019). And for the or low 30CST we utilised the guideline by Centres for Disease Control and Prevention (<11 repetitions). Notably, this physical frailty phenotype was significantly associated with lower quadriceps strength, slower walking speed, and longer 6-minute walking distance times, confirming global muscle weakness and reduced physical function in this group.

### Quality of Life (SarQoL)

The Sarcopenia Quality of Life (SarQoL) questionnaire contains 22 questions grouped into 7 domains: physical and mental health, locomotion, body composition, functionality, activities of daily living, leisure activities, and fears. Responses are recorded was self-reported, and scores were transformed to a total score ranging from 0 (lowest quality of life) to 100 (highest quality of life).

### The SARC-F (Strength, Assistance with walking, Rise from a chair, Climb stairs, and Falls)

The SARC-F questionnaire comprises five components: strength, assistance with walking, rising from a chair, climbing stairs, and falls. Each item is scored from 0 to 2, yielding a total score ranging from 0 to 10. A score of ≥4 suggests a high risk of sarcopenia and indicates the need for further diagnostic assessment. The SARC-F data was obtained via self-reported questionnaires.

### Mini Nutritional Assessment (MNA)

The MNA questionnaire consists of 18 items covering anthropometric measurements, general health status, dietary habits, and self-perceived nutritional and health status. The MNA data was obtained via self-reported questionnaires, and the total score ranges from 0 to 30, with scores ≥24 indicating normal nutritional status, 17-23.5 suggesting a risk of malnutrition, and <17 indicating malnutrition.

### Sleep Quality

Participants self-reported their sleep quality with the Insomnia Severity Index (ISI). ISI comprises seven items assessing difficulty with sleep onset, sleep maintenance, and early morning awakening, as well as dissatisfaction with current sleep, interference with daily functioning, noticeability of impairment by others, and distress caused by the sleep problem. Each item is rated on a 5-point Likert scale (0-4), yielding a total score ranging from 0 to 28. Scores are interpreted as follows: 0-7, no clinically significant insomnia; 8-14, subthreshold insomnia; 15-21, moderate clinical insomnia; and 22-28, severe clinical insomnia.

### Plasma procurement

Blood samples were collected in the morning after an overnight fast. Samples were centrifuged at 2000 g for 15-20 minutes at 4°C, aliquoted into two 2 mL microcentrifuge tubes, and stored at −80°C. Samples were then transferred to the Liverpool University Biobank Facility and were processed at the University of Liverpool for translational analysis.

### Gas Chromatography-Mass Spectrometry analysis

GC-MS was completed at the Centre for Metabolomics Research, University of Liverpool. Plasma samples were deproteinized in a single batch by adding 300 μl ice cold acetonitrile/methanol (1:1) to 100μl homogenised plasma spiked with deuterated internal standards, vortexed for 15 seconds and centrifuged at 30130 *g* for 15 minutes. Supernatant was transferred into microcentrifuge tubes (Eppendorf, Stevenage, UK) and lyophilised for 16 hours in a vacuum centrifuge (Savant SpeedVac SPD130DLX, Savant vapor trap RVT5105, (Fisher Scientific, Loughborough, UK). Pooled quality control (QC) and process blanks were prepared in tandem. Procedures for GC-MS analysis were adapted from the method presented by Dunn *et al*. (Dunn et al., 2012) Lyophilised samples, QCs and blanks were derivatized in a two-step process, randomised and analysed via autosampler injection with GC-MS. An Agilent 7250 GC-ToF-MS equipped with an HP-5ms (length 30 m, inner diameter 0.25 mm, film thickness 0.25 µm) capillary column (Agilent, Cheadle, UK) was utilised, with an oven temperature gradient of 70 °C, 4 min hold, 20 °C/min to 300 °C, 4 min hold, 300 °C transfer line temperature. MassHunter Acquisition software was set to automatically recalibrate the mass spectrometer following every four sample injections. Data were processed through spectral deconvolution, alignment, blank subtraction and retention index assignment. Peak annotation was performed using an in-house spectral library concatenated with purchased external libraries for untargeted discovery. Signal correction and quality assessment across the batch runs were carried out using pooled QC samples. The full experimental protocol available in **Supplementary Materials-Table 1**.

### Metabolomic analysis

The imputed data were log-transformed for normalization before PCA agnostic clustering. We performed agnostic clustering using principal component analysis (PCA) in MetaboAnalyst 6.0 and examined the loading plots. The top three components were identified as the most significant contributors. Hierarchical clustering was performed based on correlation of metabolites and displayed heatmaps using MetaboAnalyst 6.0. Plasma metabolites among the groups were compared, using the peak intensity data (after log transformation and normalization), with independent t-test adjusted for the Benjamini-Hochberg method for pairwise comparison and ANOVA for the Fisher’s Least Significant Difference (LSD) for 3-way comparisons.

### Statistical analysis of clinical data

Demographic, clinical, laboratory, body composition and physical function data were compared across four groups HF-NonFrail; HF-Frail; NonHF-NonFrail; NonHF-Frail. Normality testing was employed through a Shapiro-Wilk Test. Distributions across groups was compared with the Kruskal-Wallis followed by the Mann Whitney U tests and independent t-tests were used for continuous variables during the assessment of outcomes in the two groups. A chi-squared test was used for categorical variables. Least Significant Difference (LSD) test was applied for multiple comparisons, and pairwise comparison was performed using and P values were reported to show the significantly for pairwise comparison. P<0.05 determined statistical significance, using IBM SPSS version 31. Pearson correlation analysis was conducted between clinical biomarkers and physical function metrics, including HGS indexed to BMI, ALSTI indexed to BMI, total fat mass, and 30CST. Biomarkers measured included CRP, IL-6, NT-proBNP, TNF-α, activin A, GDF-15, GDF-8, follistatin-3, and the activin A to follistatin ratio. Significance was assessed at the 0.05 and 0.01 levels (2-tailed).

### Biomarker analysis

This study employed enzyme-linked immunosorbent assay (ELISA) to quantify GDF-15, myostatin, activin A, follistatin-3, tumour-necrosis-factor-α (TNF-α) [Thermo Fisher Scientific, Cat. No. KAC1751], interleukin-6 (IL-6) [Thermo Fisher Scientific, Cat. No. EH2IL6], C-reactive protein (CRP) [Thermo Fisher Scientific, Cat. No. KHA0031], glucose [Thermo Fisher Scientific, Cat. No. EIAGLUC], insulin [Thermo Fisher Scientific, Cat. No. BMS2003], and N-terminal pro-B-type natriuretic peptide (NT-proBNP) [Elabscience, Cat. No. E-EL-H6126], through commercially available human ELISA kits. Plasma samples were thawed on ice and diluted as required *as per* each biomarker’s guidelines. Standards, controls, and samples were plated in duplicates on 96-well microplates precoated with capture antibodies. After incubation with biotin conjugate, and horseradish peroxidase-conjugated streptavidin, tetramethylbenzidine substrate was added. Absorbance was measured at 450 nm (IL-6 at 450 and 540 nm) using a microplate reader. Biomarker concentrations were determined by interpolating absorbance values against standard curves generated with known concentrations. Availability of data for these biomarkers in each group are shown in **Supplementary Materials-Table 2**.

## Results

Participant characteristics are presented in **Table 1**. Between the NonHF-NonFrail NonHF-Frail, there were no significant differences in sex distribution between non-frail and frail participants (21 [11 males/10 females] vs. 8 [3 males/5 females], P= 0.68). Mean age was comparable between groups (68.4 ± 12.3 vs. 66.5 ± 8.0 years, P= 0.76). BMI tended to be higher in the NonHF-Frail group (26.5 ± 3.5 kg/m²) compared to the NonHF-NonFrail group (25.1 ± 2.8 kg/m²), although this difference did not reach statistical significance (P=0.14). Similarly, in the HF-Frail and HF-NonFrail cohort, sex distribution did not differ significantly between (7 [5 male/2 female] vs. 18 [14 male/4 female], P=0.56). Mean age was similar between HF (68.4 ± 10.7 years) and HF-Frail (67.8 ± 10.1 years) groups (P=0.89). BMI was also comparable between the two HF subgroups (31.3 ± 6.3 kg/m² vs. 30.4 ± 7.0 kg/m², P=0.77).

**Table 1.** Characteristics of the included participants. Data is presented as mean (standard deviation).

|  | **NonHF-NonFrail** | **NonHF-Frail** | **HF-NonFrail** | **HF-Frail** | ***P (among groups)*** |
| --- | --- | --- | --- | --- | --- |
| Sample (m/f) | 21 (11/10) | 8 (3/5) | 7 (5/2) | 18 (14/4) | 0.03<sup>a</sup> |
| Age (years) | 68.4 (12.3) | 66.5 (8.0) | 68.4 (10.7) | 67.8 (10.1) | NS |
| HFpEF/HFrEF | - | - | 3/4 | 14/4 | - |
| NYHA class (I-II/III) | - | - | 6/0 | 14/3 | - |
| Systolic blood pressure (mmHg) | 133.1 (13.5) | 122.5 (7.3) | 131.7 (13.7) | 120.1 (18.5) | <0.01<sup>c</sup> |
| Diastolic blood pressure (mmHg) | 80.5 (7.9) | 76.8 (7.8) | 74.3 (11.6) | 70.6 (11.9) | <0.01<sup>c</sup> |
| Resting heart rate (beats/min) | 71.0 (12.9) | 73.4 (13.1) | 59.6 (19.6) | 64.2 (10.5) | 0.048<sup>d</sup> |
| Comorbidities – n (%) |  |  |  |  |  |
| Type 2 diabetes |  |  | 2 (33.3) | 5 (29.4) |  |
| Hypertension | - | - | 3 (50.0) | 10 (58.8) | - |
| Chronic kidney disease |  |  | 1 (16.7) | 6 (35.3) |  |
| Atrial fibrillation |  |  | 3 (50.0) | 5 (29.4) |  |
| COPD |  |  | 0 (0) | 2 (11.8) |  |
| Myocardial infarction |  |  | 0 (0) | 1 (5.9) |  |
| Medications – n (%) |  |  |  |  |  |
| ACE-I/ARB |  |  | 2 (33.3) | 8 (47.1) |  |
| B-blockers | - | - | 4 (66.7) | 15 (88.2) | - |
| Loop diuretics |  |  | 3 (50.0) | 8 (47.1) |  |
| MRA |  |  | 0 (0) | 8 (47.1) |  |
| SGLT2i |  |  | 3 (50.0) | 10 (58.8) |  |
| Body mass index (kg/m<sup>2</sup>) | 25.1 (2.8) | 26.5 (3.5) | 31.3 (6.3) | 30.4 (7.0) | <0.01<sup>b</sup>,<br><0.01<sup>c</sup> |
| Fat mass (kg) | 20.83 (4.61) | 26.77 (8.28) | 33.19 (10.11) | 33.33 (14.26) | 0.047<sup>e</sup>,<br><0.01<sup>b</sup>,<br><0.01<sup>c</sup> |
| Fat free mass (kg) | 49.30 (10.49) | 45.34 (6.78) | 55.44 (11.20) | 56.69 (9.69) | 0.02<sup>c</sup>, 0.01<sup>a</sup> |
| Appendicular lean soft tissue (kg) | 20.21 (5.48) | 18.91 (3.79) | 24.00 (6.07) | 23.83 (4.43) | 0.03<sup>a</sup> |
| Appendicular lean soft tissue index (kg/m<sup>2</sup>) | 7.38 (1.04) | 6.91 (1.07) | 8.56 (1.61) | 8.04 (1.19) | 0.01<sup>b</sup>,<br>0.046<sup>c</sup>,<br>0.01<sup>d</sup>, 0.03<sup>a</sup> |
| Appendicular lean soft tissue /BMI | 0.30 (0.03) | 0.26 (0.03) | 0.28 (0.06) | 0.27 (0.03) | 0.01<sup>e</sup>, 0.03<sup>c</sup> |
| Lean soft tissue (kg) | 46.69 (9.91) | 42.84 (6.44) | 52.52 (10.60) | 53.78 (9.35) | 0.02<sup>c</sup>, 0.01<sup>a</sup> |
| Body fat (%) | 31.0 (4.9) | 37.8 (7.8) | 41.7 (12.0) | 36.9 (8.1) | <0.01<sup>e</sup>,<br><0.01<sup>b</sup>,<br><0.01<sup>c</sup> |
| Bone mineral density (g/cm<sup>2</sup>) | 1.18 (0.17) | 1.18 (0.07) | 1.23 (0.20) | 1.21 (0.12) | NS |
| Visceral adipose tissue (cm<sup>3</sup>) | 1024.1 (696.9) | 1153.1 (691.3) | 2413.3 (1847.6) | 2503.8 (1400.3) | <0.01<sup>b</sup>,<br><0.01<sup>c</sup>,<br>0.03<sup>a</sup> |
| Visceral adipose tissue (kg) | 0.97 (0.66) | 1.09 (0.65) | 2.28 (1.74) | 2.36 (1.32) | <0.01<sup>b</sup>,<br><0.01<sup>c</sup>,<br>0.03<sup>a</sup> |
| 6-minute walking distance (m) | 501.3 (79.7) | 480.3 (33.2) | 453.5 (53.9) | 386.4 (110.4) | <0.01<sup>c</sup>,<br>0.02<sup>d</sup> |
| Time up and go (seconds) | 7.36 (1.77) | 7.88 (0.84) | 8.72 (1.25) | 11.61 (4.66) | <0.01<sup>c</sup>,<br><0.01<sup>b</sup>,<br>0.04<sup>f</sup> |
| 30second chair stand (repetitions) | 14.2 (3.1) | 11.3 (1.8) | 13.9 (0.9) | 9.2 (3.1) | <0.01<sup>c</sup>,<br><0.01<sup>f</sup> |
| Gait speed (m/s) | 1.40 (0.30) | 1.26 (0.13) | 1.23 (0.22) | 1.06 (0.32) | <0.01<sup>c</sup> |
| Handgrip strength (kg) | 35.6 (9.4) | 30.3 (10.0) | 37.0 (12.2) | 30.7 (12.0) | NS |
| Handgrip strength/BMI | 1.41 (0.32) | 1.15 (0.36) | 1.22 (0.47) | 1.05 (0.45) | 0.035<sup>c</sup> |
| Knee extension strength (kg) | 36.5 (16.1) | 30.7 (16.0) | 32.0 (13.1) | 30.3 (14.1) | NS |
| Sedentary time (min/d) | 309.4 (144.8) | 377.1 (102.3) | 285.0 (158.5) | 416.0 (151.2) | 0.045<sup>c</sup> |
| Moderate activity (min/week) | 60.7 (36.5) | 40.0 (36.3) | 94.3 (127.8) | 17.2 (42.4) | 0.02<sup>c</sup> |
|  |  |  |  |  | <0.01<sup>f</sup> |
| Vigorous activity (min/week) | 37.2 (25.4) | 41.9 (81.9) | 42.9 (113.4) | 1.1 (4.7) | <0.01<sup>c</sup> |
| Metabolic equivalent of task score | 3306.1 (1970.1) | 2442.8 (3481.6) | 3531.0 (2752.4) | 678.6 (684.0) | <0.01<sup>e</sup>,<br><0.01<sup>c</sup>,<br>0.03<sup>d</sup>,<br><0.01<sup>f</sup> |
| Mini nutritional assessment score | 13.3 (1.2) | 12.4 (2.1) | 13.0 (1.0) | 10.6 (2.3) | <0.01<sup>c</sup>,<br>0.04<sup>a</sup>,<br><0.01<sup>f</sup> |
| SARC-F | 0.3 (0.6) | 1.0 (1.4) | 1.7 (1.7) | 2.9 (3.5) | <0.01<sup>c</sup> |
| SarQoL | 83.7 (10.0) | 75.5 (16.6) | 65.8 (12.8) | 57.9 (17.6) | 0.01<sup>b</sup>,<br><0.01<sup>c</sup>,<br><0.01<sup>a</sup> |
| Insomnia severity index | 3.6 (3.2) | 5.4 (6.7) | 7.0 (4.2) | 10.8 (8.2) | <0.01<sup>c</sup> |
| Insulin (pg/mL) | 656.0 (807.1) | 462.9 (343.1) | 287.1 (200.4) | 818.9 (1126.0) | NS |
| Glucose (mg/dL) | 85.2 (15.8) | 79.6 (10.6) | 93.0 (17.3) | 111.8 (60.9) | 0.01<sup>a</sup><br>0.02<sup>d</sup> |
| HOMA-IR | 2.58 (3.87) | 1.61 (1.33) | 1.22 (1.14) | 6.22 (14.90) | <0.01<sup>b</sup> |
| CRP (mg/dL) | 1.92 (2.19) | 1.98 (1.26) | 2.99 (3.03) | 2.85 (2.83) | NS |
| IL-6 (pg/mL) | 65.5 (127.7) | - | 3.45 (4.40) | 45.2 (128.3) | NS |
| NT-proBNP (pg/mL) | 117.1 (114.2) | 111.1 (88.2) | 104.1 (25.0) | 241.2 (198.0) | 0.01<sup>c</sup> |
| TNF-a (pg/mL) | 4.44 (1.63) | 4.43 (1.50) | 8.45 (7.00) | 7.04 (2.24) | 0.01<sup>a</sup><br>0.007<sup>c</sup> |
| Follistatin-3 (pg/mL) | 10,531.4 (15,288.1) | 25,807.4 (56,244.5) | 36,712.8 (36,819.8) | 33,432.8 (53,446.3) | NS |
| Activin A (pg/mL) | 594.9 (719.4) | 486.3 (386.9) | 213.4 (161.3) | 155.3 (143.4) | 0.046<sup>a</sup> |
| Activin A/Follistatin-3 | 0.16 | 0.15 | 0.01 | 0.03 | 0.037<sup>c</sup> |
| GDF-15 (pg/mL) | 382.5 (236.0) | 556.3 (391.8) | 456.4 (395.8) | 1227.2 (933.2) | <0.01<sup>a</sup><br>0.03<sup>c</sup> |
| Myostatin (ng/mL) | 1.15 (0.65) | 1.32 (1.59) | 1.15 (0.81) | 1.45 (1.56) | NS |

The HF-Frail had the significantly lower self-reported moderate physical activity compared to HF-NonFrail (P<0.01) and NonHF-NonFrail (P = 0.04). The self-reported MNA score, and SarQoL was lower HF-Frail (P<0.01), compared to HF-NonFrail, and NonHF-NonFrail. HF-Frail had the highest BMI (P<0.01), followed by HF-NonFrail (P<0.01), and NonHF. Whereas HF-Frail had the lowest appendicular lean soft tissue/BMI (P=0.03), followed by HF-NonFrail and NonHF-NonFrail. The HF-Frail compared to NonHF-NonFrail had consistently lower physical function as indicated with longer 6-minute walking distance times (P<0.01), slower time Up and Go (P<0.01), lower repetitions in 30 Chair Stand Test (P<0.01), and weaker hand grip strength (P=0.035). HF-Frail compared to NonHF-NonFrail had elevated plasma levels for GDF-15 (P=0.03), NT-proBNP (P=0.01), and TNF-a (P=0.007).

### Metabolic alterations in total HF compared to total NonHF

A total of 149 unique metabolites and 296 unidentified features were detected. The m/z values, retention times, and p-values obtained from independent t-tests were used as input for Mummichog functional analysis (Table 2). This approach mapped 22 metabolic pathways, of which the most significantly enriched (p < 0.05) were beta-alanine metabolism; valine, leucine, and isoleucine degradation; C21-steroid hormone biosynthesis and metabolism; vitamin A (retinol) metabolism; urea cycle and amino group metabolism; and androgen and estrogen biosynthesis and metabolism.

**Table 2.** Mummichog pathway enrichment. Analysis for HF and nonfailing control based on top 10% significant differences.

|  | Pathway total | Hits.total | Hits.sig | P(Fisher) | P(EASE) | P(Gamma) |
| --- | --- | --- | --- | --- | --- | --- |
| Beta-Alanine metabolism | 20 | 6 | 3 | 0.059847 | 0.25195 | 0.00449 |
| Valine, leucine and isoleucine degradation | 65 | 7 | 3 | 0.092707 | 0.31801 | 0.0055708 |
| C21-steroid hormone biosynthesis and metabolism | 112 | 8 | 3 | 0.13142 | 0.38285 | 0.0069202 |
| Vitamin A (retinol) metabolism | 67 | 3 | 2 | 0.072875 | 0.41382 | 0.0076915 |
| Urea cycle/amino group metabolism | 85 | 17 | 4 | 0.3081 | 0.54342 | 0.012193 |
| Androgen and estrogen biosynthesis and metabolism | 95 | 5 | 2 | 0.19484 | 0.59116 | 0.014586 |
| Aspartate and asparagine metabolism | 114 | 28 | 4 | 0.72558 | 0.87186 | 0.052562 |
| Tryptophan metabolism | 94 | 23 | 2 | 0.92603 | 0.98645 | 0.15467 |
| Tyrosine metabolism | 160 | 23 | 2 | 0.92603 | 0.98645 | 0.15467 |
| Histidine metabolism | 33 | 4 | 1 | 0.52024 | 1 | 1 |
| Vitamin B5 - CoA biosynthesis from pantothenate | 12 | 2 | 1 | 0.30615 | 1 | 1 |
| Bile acid biosynthesis | 82 | 3 | 1 | 0.42279 | 1 | 1 |
| Biopterin metabolism | 22 | 3 | 1 | 0.42279 | 1 | 1 |
| Glycine, serine, alanine and threonine metabolism | 88 | 11 | 1 | 0.8718 | 1 | 1 |
| Fatty Acid Metabolism | 63 | 6 | 1 | 0.66944 | 1 | 1 |
| CoA Catabolism | 7 | 3 | 1 | 0.42279 | 1 | 1 |
| Butanoate metabolism | 34 | 6 | 1 | 0.66944 | 1 | 1 |
| Arginine and Proline Metabolism | 45 | 13 | 1 | 0.9128 | 1 | 1 |
| Glutamate metabolism | 15 | 1 | 1 | 0.16667 | 1 | 1 |
| Benzoate degradation via CoA ligation | 4 | 1 | 1 | 0.16667 | 1 | 1 |
| Pyrimidine metabolism | 70 | 4 | 1 | 0.52024 | 1 | 1 |
| Lysine metabolism | 52 | 13 | 1 | 0.9128 | 1 | 1 |

Figure 1A shows the principal component analysis (PCA), revealing separable clusters between the whole HF (HF-Frail+ HF-NonFrail) and Non-HF (NonHF-Frail & NonHF-NonFrail) groups, with results presented in **Table S1**. In contrast, HFpEF and HFrEF had substantial overlap (Figure 1B). Several fatty acid related metabolites showed altered levels in total HF. Octadecadienoic acid (P<0.001), octadecenoic acid methyl ester (P<0.001), and octadecenoic acid (P=0.037) were significantly higher in total. Also, tryptophan (P=0.002) and indole-3-acetamide (P=0.044) were higher in total Non-HF controls, while indole-3-hydroxy (P=0.0873) and indole-3-acetic acid (P=0.3618) trended higher in HF. Several branched-chain amino acids (BCAAs) and related metabolites were elevated in HF, such as isoleucine (P<0.001), valine (P=0.00598), and ornithine (P<0.001). Branched-chain fatty acid derivatives including isovaleric acid 2-oxo (P<0.001) and valeric acid 2-oxo (P=0.0256) were also higher in HF. Regarding other amino acids, alanine (P<0.001) aspartic acid (P=0.00293), glutamic acid (P<0.001), cystine (P=0.00107), and histidine (P<0.001) were significantly different between groups, with negative t-values indicating higher abundance in HF. In contrast, glutamine (P=0.719), asparagine (P<0.001), serine (P<0.001and methionine (P<0.001) showed strong group differences, with asparagine and serine elevated in total Non-HF controls and methionine reduced in HF. Several glycolysis and glycolysis-related metabolites were altered. Glucose 2-deoxy (P=0.0232), glycerol (P<0.001), galactose (P<0.001), galacturonic acid (P< 0.001), and GalNAcol (P<0.001) were increased in HF, whereas fructose (P=0.00115) and gluconic acid-1,4-lactone (P<0.001) were reduced. Additionally, several metabolites showed significant differences, such as galactonic acid (P<0.001), and cystamine (P<0.001), between total HF and total Non-HF. Results of hierarchical clustering also confirms the separation between total HF and total NonHF as illustrated in Figure 1C.

**Figure 1.**
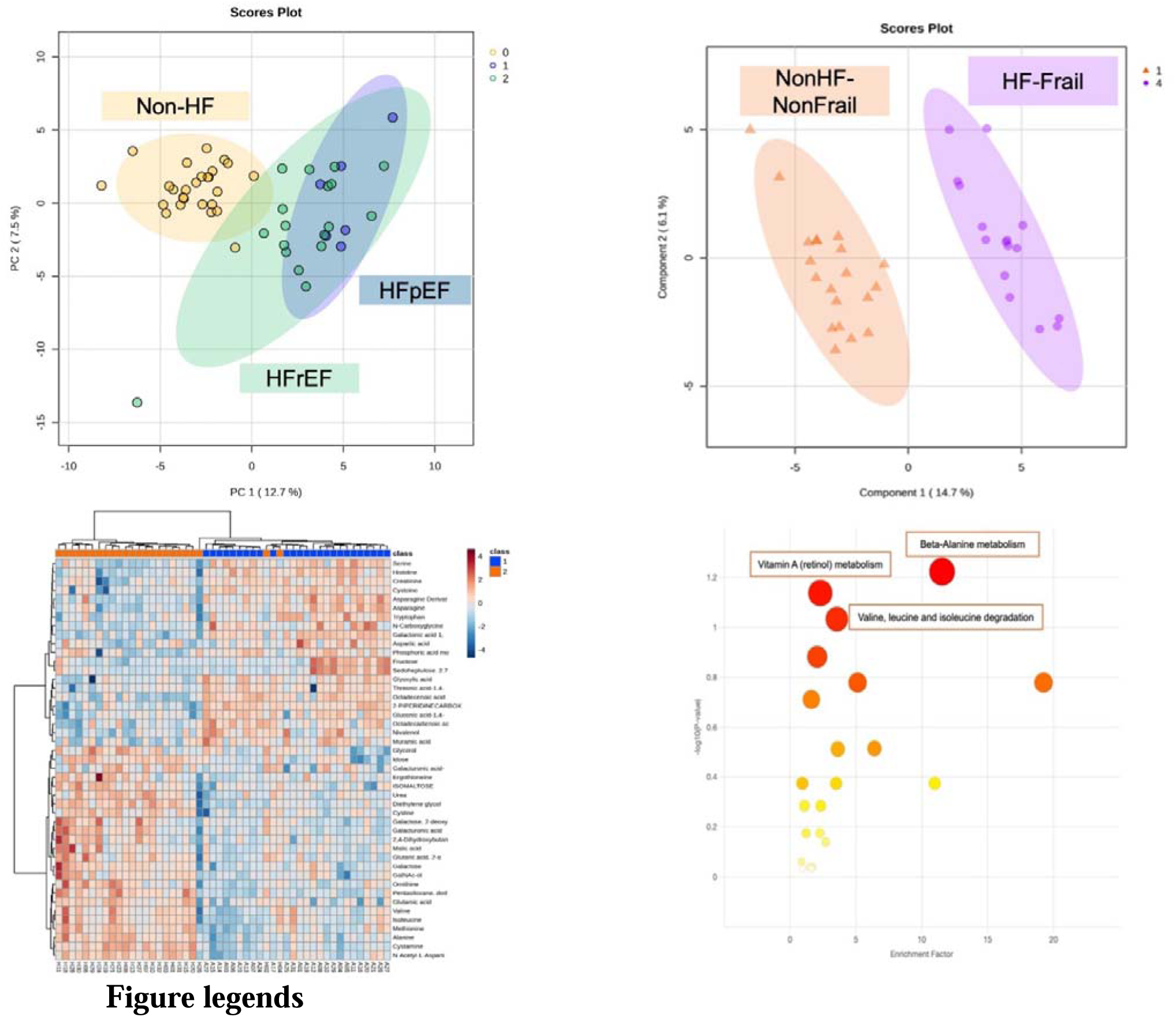
Plasma metabolomic signatures in HF and nonfailing controls. **A**, PCA analysis shows distinct clusters for HF compared to nonfailing controls [F-value: 44.636; R-squared: 0.4767; p-value (based on 999 permutations): 0.001], **B**, and substantial overlap between HFrEF and HFpEF. **C**, Hierarchical clustering of participants (columns) using the plasma metabolome demonstrates similar separation of the HF group from nonfailing control. **D**, Mummichog Pathway Activity Profile for HF compared to non-failing controls based on top 10% peaks.

We also performed mummichog analysis as illustrated in Figure 1D and data presented in **Table S2.** Metabolic pathway enrichment analysis identified several significantly affected pathways. The most prominent was beta-alanine metabolism (P Gamma=0.0054), followed by urea cycle/amino group metabolism (P for Gamma=0.0072), Lysine metabolism (P for Gamma=0.0087), and Aspartate and asparagine metabolism (P for Gamma=0.0172), and valine, leucine and isoleucine degradation (P for Gamma=0.0305), indicating disruptions in branched-chain amino acid catabolism.

### Metabolic alterations accounting for frailty phenotype in HF

In pairwise comparisons between HF-Frail and NonHF-NonFrail (**Table S2**), several metabolites were significantly higher in HF-Frail (P< 0.05), including cystamine (P<0.001), alanine (P<0.001), N-acetyl-L-aspartic acid (P<0.001), methionine (P<0.001), and galactose (P<0.001). Higher in controls were 2-piperidinecarboxylic acid (P<0.001) galactonic acid 1,4-lactone (P<0.001), octadecenoic acid methyl ester (P<0.001), N-carboxyglycine (P<0.001), and histidine (P<0.001). Differences were also observed in tricarboxylic acid (TCA) cycle intermediates such as malic acid (P<0.001), fumaric acid (P=0.008) and Glutaric acid. 2-oxo (P<0.001). Branched-chain amino acids (BCAA) including isoleucine (P=0.0174) and valine (P=0.003) along with related catabolites, indicating broad coverage of core metabolic pathways in the observed alterations.

We screened metabolites in HF-Frail compared to those with HF but without frailty. The PCA results did not yield a significant model. Further, the supervised (PLS-DA) analyses were applied to select the variables responsible for the separation showed by models in Figure 2. The two first components showed the highest accuracy which were used as loading plots to generate the variable of Importance (VIP>1.0) contributing to the model. Differing VIPs between HF-Frail compared to their counterparts based on raw P-values included lower galacturonic acid-1-phosphate (P=0.03), glutamic acid (P=0.04), indole-3-acetamide (P<0.01), methionine (P=0.04), and phenylalanine P=0.04). On the contrary, higher amounts of 2-hydroxy-glutaric acid were observed (P=0.02) (**Table S3**).

**Figure 2.**
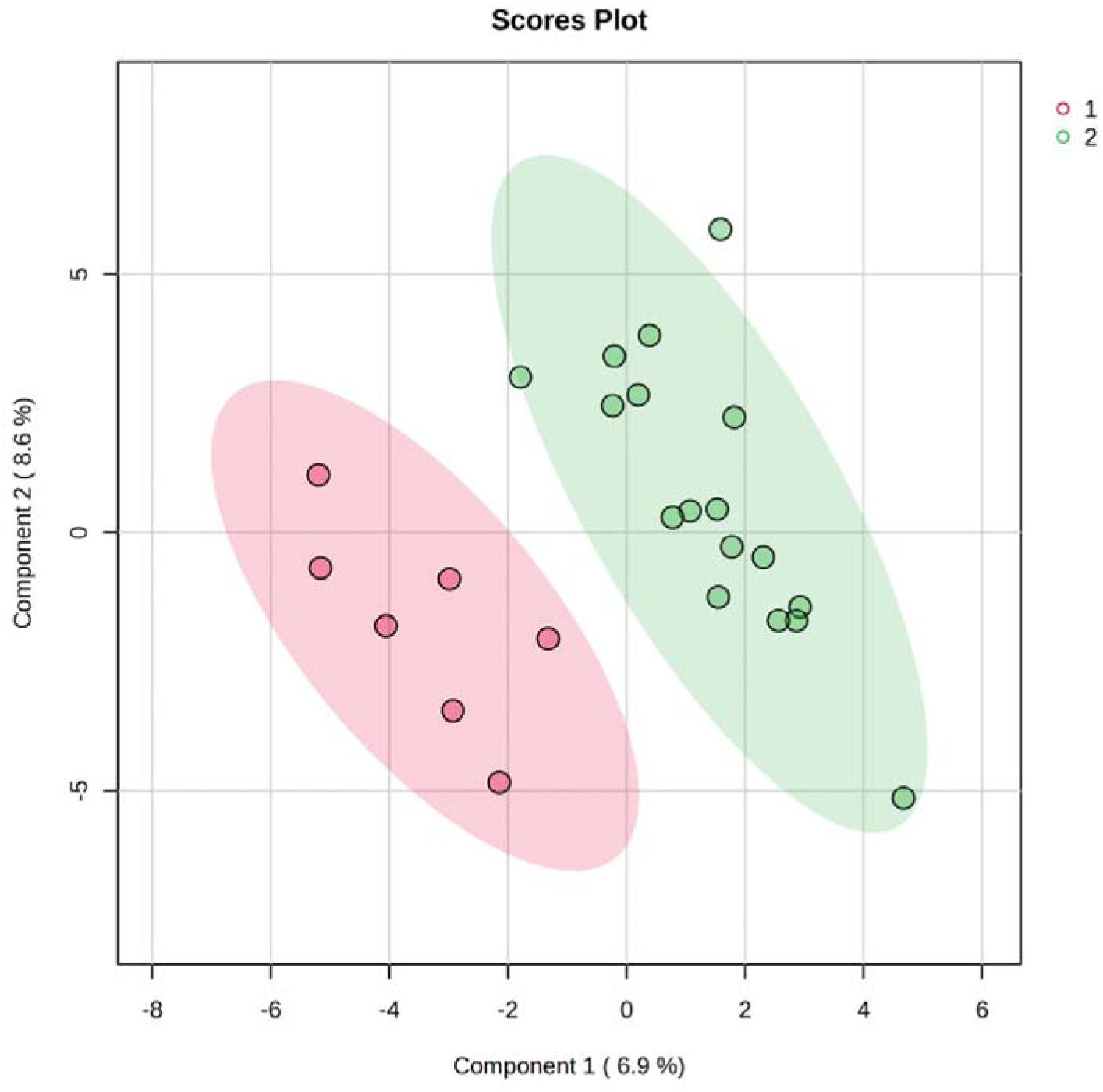
Scor e plots of parti al least squar es discr imin ant analy sis (PLS-DA) performed on gas chromatography–mass spectrometry data of plasma samples collected from HF-Frail and HF-NonFrail.

### Correlations between physical function and body composition with biomarkers

Significant negative correlations were observed: HGS/BMI with NT-proBNP (r = −0.41, P=0.01) and GDF-15 (r = −0.346, P=0.02); ALSTI/BMI with TNF-α (r = −0.304, P=0.04); total fat mass with TNF-α (r = −0.414, P>0.01) and activin A to follistatin-3 ratio (r = −0.350, P=0.04); and 30CST with NT-proBNP (r = −0.433, P<0.01) and GDF-15 (r = −0.634, P<0.01) (**Table 3**).

**Table 3.**
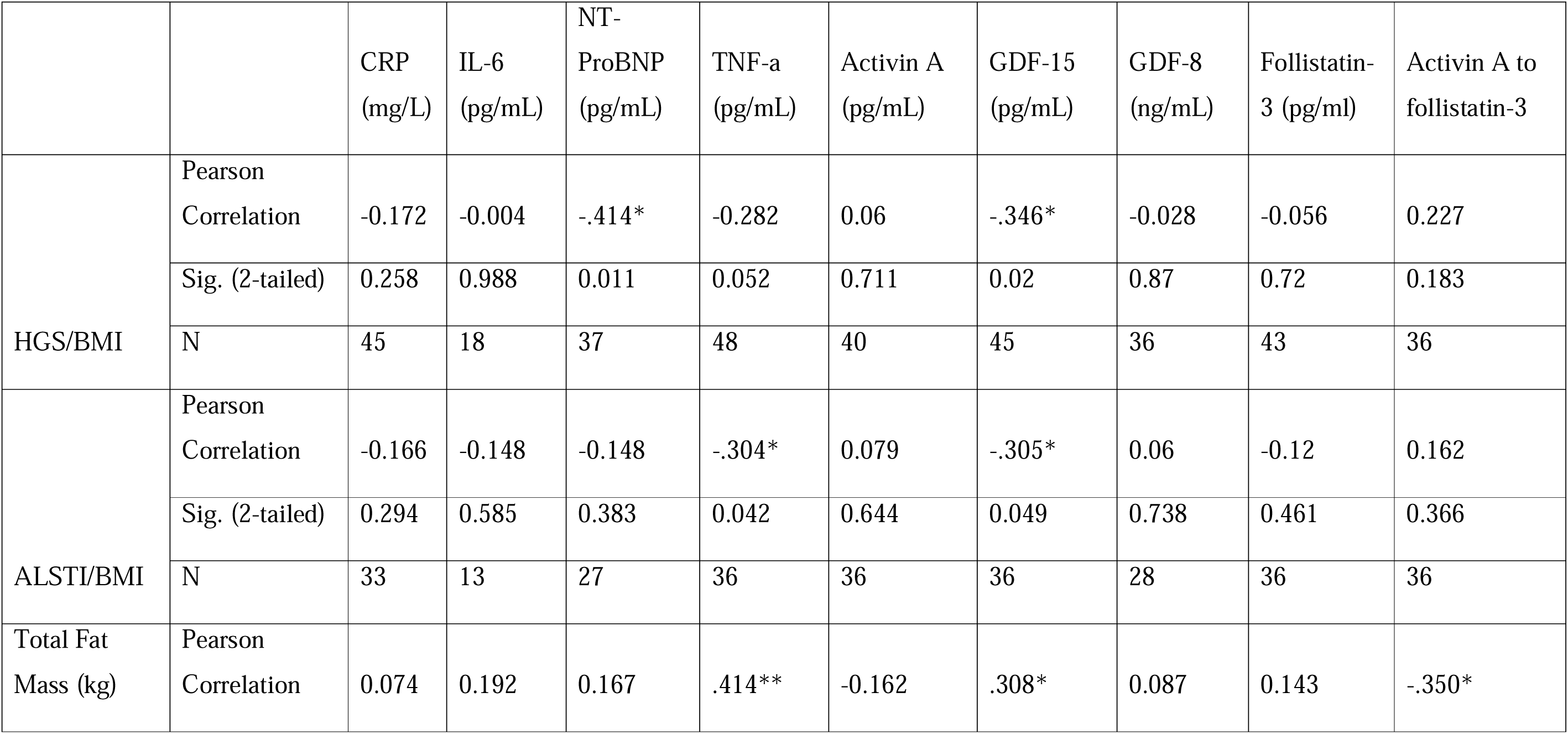
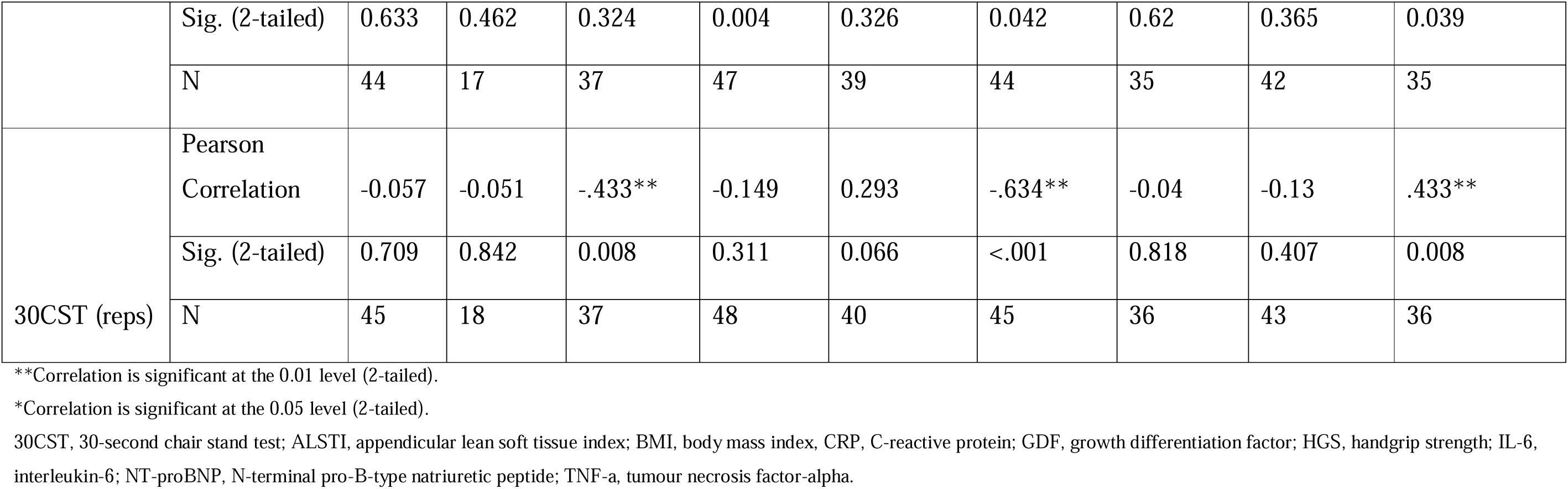
Pearson correlation for clinical biomarkers, with handgrip strength indexed to BMI, ALSTI indexed to BMI, total fat mass and 30CST.

| | | CRP<br>(mg/L) | IL-6<br>(pg/mL) | NT-ProBNP<br>(pg/mL) | TNF-α<br>(pg/mL) | Activin A<br>(pg/mL) | GDF-15<br>(pg/mL) | GDF-8<br>(ng/mL) | Follistatin-3 (pg/ml) | Activin A to<br>follistatin-3 |
| --- | --- | --- | --- | --- | --- | --- | --- | --- | --- | --- |
| HGS/BMI | Pearson<br>Correlation | -0.172 | -0.004 | -.414* | -0.282 | 0.06 | -.346* | -0.028 | -0.056 | 0.227 |
|  | Sig. (2-tailed) | 0.258 | 0.988 | 0.011 | 0.052 | 0.711 | 0.02 | 0.87 | 0.72 | 0.183 |
|  | N | 45 | 18 | 37 | 48 | 40 | 45 | 36 | 43 | 36 |
| ALSTI/BMI | Pearson<br>Correlation | -0.166 | -0.148 | -0.148 | -.304* | 0.079 | -.305* | 0.06 | -0.12 | 0.162 |
|  | Sig. (2-tailed) | 0.294 | 0.585 | 0.383 | 0.042 | 0.644 | 0.049 | 0.738 | 0.461 | 0.366 |
|  | N | 33 | 13 | 27 | 36 | 36 | 36 | 28 | 36 | 36 |
| Total Fat<br>Mass (kg) | Pearson<br>Correlation | 0.074 | 0.192 | 0.167 | .414** | -0.162 | .308* | 0.087 | 0.143 | -.350* |
|  | Sig. (2-tailed) | 0.633 | 0.462 | 0.324 | 0.004 | 0.326 | 0.042 | 0.62 | 0.365 | 0.039 |
|  | N | 44 | 17 | 37 | 47 | 39 | 44 | 35 | 42 | 35 |
| 30CST (reps) | Pearson<br>Correlation | -0.057 | -0.051 | -.433** | -0.149 | 0.293 | -.634** | -0.04 | -0.13 | .433** |
|  | Sig. (2-tailed) | 0.709 | 0.842 | 0.008 | 0.311 | 0.066 | <.001 | 0.818 | 0.407 | 0.008 |
|  | N | 45 | 18 | 37 | 48 | 40 | 45 | 36 | 43 | 36 |

\*\*Correlation is significant at the 0.01 level (2-tailed).
\*Correlation is significant at the 0.05 level (2-tailed).
30CST, 30-second chair stand test; ALSTI, appendicular lean soft tissue index; BMI, body mass index, CRP, C-reactive protein; GDF, growth differentiation factor; HGS, handgrip strength; IL-6, interleukin-6; NT-proBNP, N-terminal pro-B-type natriuretic peptide; TNF-a, tumour necrosis factor-alpha.

## Discussion

In this study, we have shown that low physical activity alongside with lower 30CST (lower extremity strength) and low GS (upper extremity strength) can show a clinically meaningful distinction of frailty indices, and was associated with lower quality of life, and higher malnutrition. Levels of TNF-α, and GDF-15 was elevated in HF-Frail followed by HF-NonFrail, while the GDF-15 seems to have to most distinctive values when compared to NonHF-NonFrail (Figure 3). Further, metabolomics analysis revealed disruptions in HF-NonFrail and HF-Frail across the fatty acid-related metabolites, amino acid metabolism, and glycolytic pathways compared to NonHF controls, indicating impaired energy production and a catabolic state. Finally, we found significant negative correlations between NT-proBNP, TNF-α, GDF-15, suggesting that markers of inflammation and cardiac stress may be linked to reduced muscle quality and physical function. These observations display new and interconnected metabolic links that may implicate pathobiological mechanisms driving physical frailty and muscle weakness in HF.

**Figure 3.**
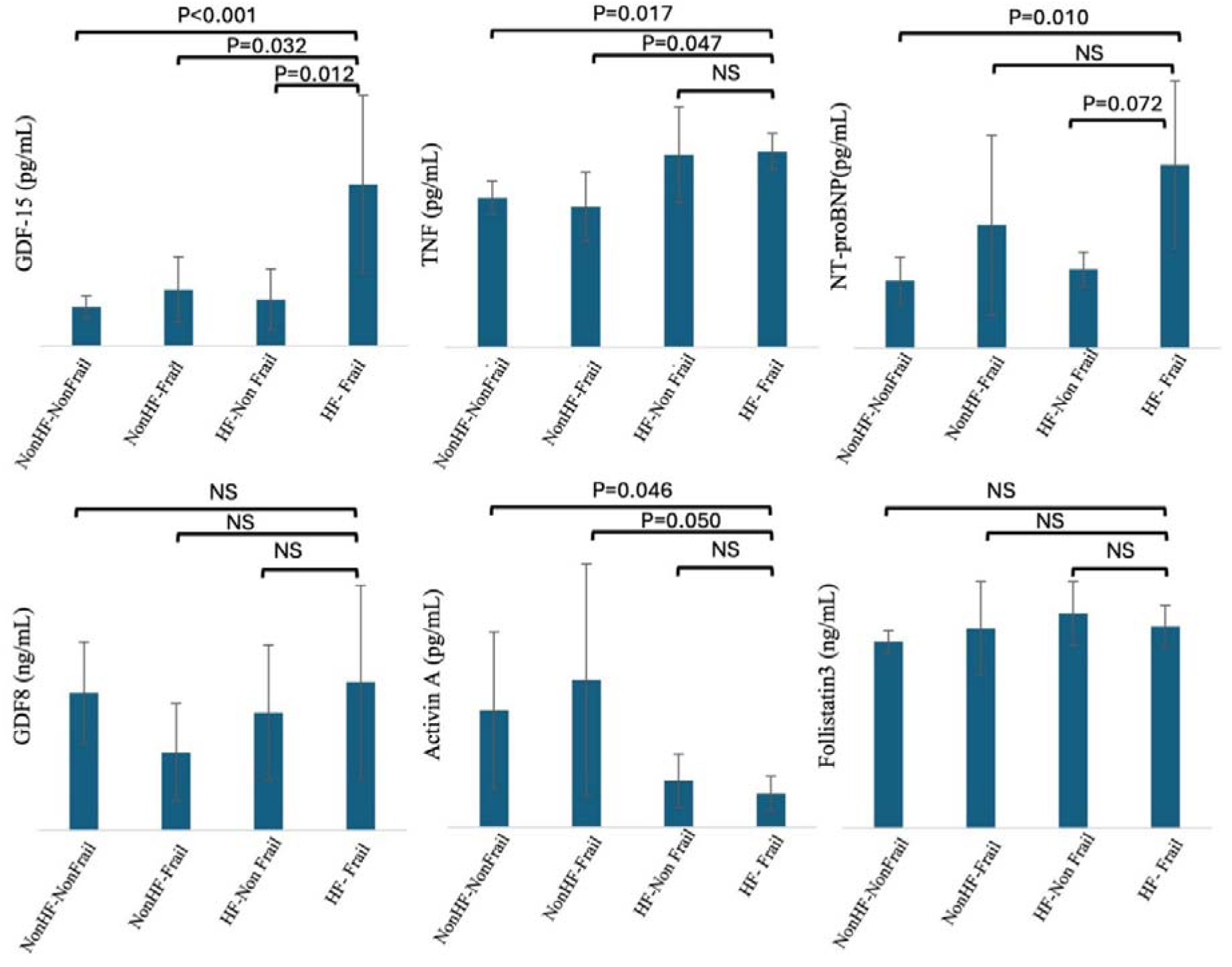
Biomarker levels are displayed as the geometric mean with a corresponding 95% CI of the X shows the mean. A) Displays the differences in plasma GDF-15 (growth differentiation factor-15) observing higher levels in HF-frail vs. HF-Not frail (P=0.02), and NonHF-Frail (P<0.01), and NonHF-Not frail (P=0.03). B) HF-Frail had higher level of TNF-a compared to NonHF-NonFrail (P=0.007) and NonHF-Frail (P=0.01). C) NonHF-Not frail displayed lower NT-proBNP levels (117.1 (114.2) pg/mL) than HF-Frail (241.2 (198.0) pg/mL; P=0.01). D) HF-Frail had higher lowest level of Activin A/ Follistatin-3 (P=0.037) compared to other groups.

### Metabolic alterations in HF compared to NonHF controls

We initially investigated the plasma global metabolomic profiles distinguished the outcomes of interest total HF and total Non-HF and then followed this for HF-Frail and NonHF-NonFrail. Principal component analysis showed clear patterns of separation, which is notable in our chronic-HF phenotype (Figure 1). Elevated levels of hexadecenoic acid and BCAAs such as isoleucine and valine, along with their derivatives (isovaleric acid 2-oxo and valeric acid 2-oxo), were observed in HF. These findings align with prior research indicating that HF is associated with disrupted lipid and amino acid metabolism, likely due to impaired energy utilization and increased catabolic demands (Lopaschuk et al., 2021). Elevation of BCAAs in HF may reflect impaired mitochondrial oxidative capacity via increased reactive oxygen species production and dysregulation of mitochondrial Ca^2+^ homeostasis (Smith and Eisner, 2019), increased risk of insulin resistance (Wang et al., 2011), and disrupted cardiac energy substrate utilization, albeit a small amount of BCAAs is used in cardiac proteolysis (Murashige et al., 2020). Conversely, reductions in glutamine, asparagine, serine, and methionine in patients with HF suggest increased protein turnover and inefficient adenosine triphosphate (ATP) production to meet energy demands, for which, previous research has shown links to BNP, left ventricular ejection fraction and end-diastolic volume index, and inferior vena cava diameter (Hakuno et al., 2015), potentiating risks of incident myocardial infarction. In addition, the observed increases in glycolytic metabolites, such as glucose-2-deoxy, glycerol, galactose, galacturonic acid, and galNac-ol, and decreased fructose and gluconic acid, highlight altered carbohydrate metabolism in HF, which have been linked with cardiomyopathies and altered glucose transporter regulation (GLUT1 and GLUT4), that facilitate cardiac contractile function (Tran and Wang, 2019). Overall, these changes may reflect compensatory mechanisms to sustain energy production in the failing heart, where oxidative phosphorylation is compromised. The elevation of glucose-related metabolites may also indicate insulin resistance, as supported by the higher HOMA-IR scores in patients with HF-Frail.

### Metabolic alterations in heart failure with frailty

Our findings regarding the HF-Frail phenotype revealed several differentially expressed metabolites, including galacturonic acid-1-phosphate, glutamic acid, indole-3-acetamide, methionine, phenylalanine, and 2-hydroxy-glutaric acid. These findings, although based on raw P-values, suggest that frailty in HF may amplify metabolic perturbations compared to patients with HF but without frailty. However, the elevation of amino acids such as glutamic acid and phenylalanine in HF-Frail yields controversial results regarding their link to frailty (Shekarchian et al., 2024, Carbone et al., 2024, Zhou et al., 2024). Regarding HF, higher phenylalanine has been linked to increased mortality and HF (re)hospitalization (Chen et al., 2020, Wang et al., 2019), while glutamic acid may reflect HF severity (Yang et al., 2024), that may be exacerbated by frailty. Furthermore, the higher levels of galacturonic acid-1-phosphate, a carbohydrate metabolite, may reflect altered glycosaminoglycan metabolism, potentially contributing to impaired tissue remodelling, considering that proteomics have demonstrated it as a top pathway associated with frailty (Sathyan et al., 2020). It is worth noting that the HF-Frail cohort exhibited lower 30CST performance, reduced weekly moderate physical activity, and lower MET and MNA scores compared to patients with HF but without frailty.

These changes were accompanied by higher levels of glucose, TNF-α, and GDF-15, but lower levels of activin A compared to HF without frailty. Elevated GDF-15 is a biomarker of inflammation and oxidative stress that has been previously linked to sarcopenia and frailty (Kamper et al., 2024). This suggests that GDF-15 may not only serve as a marker of HF hospitalization and mortality (Teramoto et al., 2024), or inflammation and malnutrition (Sakamoto et al., 2025), but also of frailty and/or myopathy severity in HF. In addition, higher activin A has been linked to impaired cardiac remodelling (Yndestad et al., 2004), and myocardial damage (Oshima et al., 2009) in humans and murine models, which contradict our results on the detrimental impact physical frailty may confer in HF. Our data showed that the ratio of activin A to follistatin-3 was lower in both the HF and HF-Frail groups compared to controls, suggesting that follistatin-3, as a natural antagonist, may sequester activin A more effectively in the HF-Frail phenotype.

The elevated levels of TNF-a in our study are in line with evidence on its link with frailty in cardiovascular diseases (James et al., 2024). Overall, HF-Frail is characterized by altered galacturonic acid-1-phosphate, amino acids, glucose, TNF-α, and GDF-15, but lower activin A, reflecting increased metabolic stress, inflammation, and impaired tissue remodelling compared to HF counterparts without frailty. This observation suggests a multifactorial yet tightly interconnected relationship between heart failure (HF) and a muscle catabolic state. Preclinical models have demonstrated that insulin resistance and impaired mitochondrial respiratory capacity in skeletal muscle are associated with marked increases in GDF-15 expression and elevated plasma GDF-15 concentrations, indicating that skeletal muscle may be a primary source of increased circulating GDF-15 observed in middle-aged mice (Chen et al., 2025).

HF-Frail exhibited a broad range of metabolic alterations compared to NonHF controls without frailty. For instance, elevated amino acids (e.g., alanine, methionine, isoleucine, glutamic acid, ornithine, and cystine) and carbohydrate metabolites (e.g., galactose, galactonic acid, and glycerolaldopyranosid) suggest increased metabolic stress. Conversely, reduced levels of energy metabolism intermediates (e.g., pyruvic acid, malic acid, and 2-oxo-glutaric acid) indicate compromised mitochondrial energy production, a feature consistent with the systemic impact of frailty on energy homeostasis, which was also observed in the total HF vs. NonHF cohorts. The lower concentrations of tryptophan, histidine, serine, cysteine, and asparagine further highlight the catabolic state in HF-Frail, linking these to potentially accelerated muscle wasting and reduced production of serotonin, kynurenines, and NAD^+^ as shown previously (Migliavacca et al., 2019). Thus, patients with HF and frailty may exhibit extensive metabolic perturbations, including elevated amino acids and carbohydrate metabolites, and reduced energy intermediates compared to NonHF controls without frailty.

### Link between muscle quality and biomarkers

The observed negative correlations between NT-proBNP, TNF-α, GDF-15, and indices of sarcopenia and body composition, such as HGS/BMI, ALSTI/BMI, total fat mass, and 30CST, indicate a potential link between elevated inflammatory and cardiac stress markers and diminished physical function. Notably, the strong association of GDF-15 with 30CST (r = −0.634) may be a key indicator of functional decline and sarcopenia risk as previously observed in the PROTECT cohort consisting of older medical patients (Kamper et al., 2024). These findings align with the hypothesis that chronic inflammation and cardiac stress may share a bidirectional relationship with muscle weakness. However, the cross-sectional nature of the data limits causal inference, warranting longitudinal studies to elucidate these correlations.

### Clinical and research implications

The notable muscle weakness in HF-Frail was apparent at the similar age with NonHF and HF counterparts, suggesting that physical frailty can start at younger age in this population, which is an established risk factor for worsened prognosis in HF. The distinct metabolic profiles identified in this study could have significant implications for the management of HF, particularly in the context of physical frailty. The elevation of GDF-15 and TNF-α in HF-Frail highlights the role of inflammation, and oxidative stress in this population, suggesting that potential therapies (pharmacological and non-pharmacological) could be explored to mitigate frailty-related complications. Notably, GDF-15 has recently emerged as a therapeutic target in cancer-associated cachexia, a condition characterized by severe muscle and weight loss (Groarke et al., 2024). Additionally, the observed metabolic alterations, particularly in BCAAs, glycolytic metabolites, and the tryptophan-indole pathway, suggest potential therapeutic targets to improve energy metabolism and muscle health in HF-Frail.

For example, interventions aimed at enhancing mitochondrial function or optimizing amino acid metabolism could help alleviate frailty symptoms and improve functional outcomes (Buondonno et al., 2020, Mishra et al., 2024). Specifically, nutritional status, as indicated by lower MNA scores in HF with frailty, may be a critical factor, given its link to lower energy and protein intake, amplifying sarcopenia risk. Additionally, the lower 30CST performance and physical activity levels in the HF-Frail group, marking further the need for tailored nutritional and exercise programs to improve physical function(Mirkowski et al., 2025).

From a research perspective, the study highlights the importance of integrating frailty assessments into studies with participants who have HF to better understand its metabolic contributions. The lack of a significant model when screening metabolites between HF with and without frailty suggests that larger sample sizes or more sensitive analytical approaches may be needed in studies to fully elucidate frailty-specific metabolic signatures. Future studies could employ longitudinal designs to explore how these metabolic changes evolve over time and evaluate their prognostic impact on clinical outcomes such as hospitalization or mortality.

### Study limitations

The relatively small sample size and cross-sectional nature precludes causal interpretations and to investigate the physical frailty bidirectionally to HF, particularly for frailty-specific analyses, where statistical significance was based on raw p-values rather than adjusted models. Additionally, the consideration of comorbidities and different prescribed medications may have an impact on metabolomic analyses. MSI level 2 annotations, not definitive metabolite identifications. Future work would be to run authentic standards to confirm the ones not already in CMR library, although hits in mummichog give extra confidence. Despite these constraints, the robust application of GC-MS in clinical and community-dwelling cohorts, provides further mechanistic insights into HF and physical frailty. These findings highlight the need for integrated approaches to manage HF and frailty, incorporating nutritional, exercise, and potentially pharmacological interventions to address metabolic and functional deficits.

## Conclusions

This study provides novel insights into the metabolic alterations associated with HF and frailty, highlighting distinct profiles in lipid, amino acid, and carbohydrate metabolism. HF-Frail patients demonstrate exacerbated impaired energy metabolism, marked by reduced ability to replenish excess tricarboxylic acids intermediates, and altered amino acid and carbohydrate metabolism. Higher BCAAs and its catabolites, at least partly, indicate high systemic catabolism, alongside elevated inflammation and GDF-15, are markers of muscle wasting.

## Supporting information

Supplementary Materials alongside Table S1-S3

## Acknowledgements

The authors would like to express their sincere gratitude to all participants of this study. We also gratefully acknowledge the invaluable contributions of the Liverpool University Biobank team, Dr. Viktoria Shaw, Mrs. Sue Holden, and Mrs. Katie Bullock for their support in biomarker analysis, as well as the processing and storage of samples and data used in this study.

## Authors’ contributions

KP and MI conceptualized the study. KP, SMJF, BGA, and AN conducted the sarcopenia assessments. AB and HM conducted the metabolomic sample preparation and analysis. KP and MI conduced the statistical analyses. KP, SJF, BGA, and MI wrote the manuscript. MI supervised the manuscript. OK, GYHL, RS, HM, and MI revised the manuscript.

## Data availability statement

Metabolomics data coupled to deidentified metadata may be provided on reasonable request.

## Ethical approval statement

This study received ethical approval from the London-Queen Square Research Ethics Committee (REC Reference: 23/PR/0050).

## Pre-registered clinical trial number

NCT06217640

