## Supplementary Materials alongside Table S1-S3 for "Heart failure with physical frailty is associated with inflammation, insulin resistance, GDF-15 and impaired energy and amino acid metabolism"

### Study Design

**Study Locations:** Liverpool University Hospitals Foundation Trust (Clinical lead: Dr Rajiv Sankaranarayanan, Heart Failure Consultant, Aintree Heart Failure Clinic). The Liverpool University Hospitals NHS Foundation Trust (LUFHT) sees approximately 2000 HF patients annually and our recruitment target is 60 HF patients over six months period (less than 5% for a six-month period). Assessments can take place at the Hospital site, or at the Liverpool Hope University, the only difference is the DEXA scan which will be only possible at Liverpool Hope University. All participants will receive information and can choose whether to attend.

**Access to medical record:** Data from patient was obtained from medical records.

**Recruitment**: patients has been identified by reviewing the medical records and admission record, against the inclusion and exclusion criteria by a member of the research team and have been contacted to ask for their interests to take part in the study. Those willing to take part have received the consent form, patient information sheet, and questionnaires prior to visit. After the initial explanation, the participant would have 2 days to discuss this with friend, carer, or family member. Once informed consent has been obtained, by post or in person basic demographics, medical, drug and social history would be recorded from the patient’s medical notes. An assessment visit day then will be scheduled. Participant received compensation according to NIHR protocol.

### Body Composition Assessment

Whole-body composition was measured using whole-body GE Lunar iDXA scan (GE Healthcare, Madison, WI, USA), following manufacturers standard operating procedures. Participants were positioned supine on the scanning bed, wearing light clothing and no metal items to ensure measurement accuracy. The total body scan mode was used to measure total mass, lean mass, and fat mass. DXA quality control procedures were conducted according to the manufacturer's guidelines, and daily calibration was performed to ensure measurement accuracy. Following the scan, the system’s latest software release (enCore version 18) was used to process and analyse the body composition results.

### Physical Function Assessment

**Handgrip strength** of dominant and non-dominant sides was measured using a portable Jamar hand grip dynamometer (Biometric Ltd, Wireless Dynamometer G200, Newport, UK). Participants sat upright in an armless chair, with shoulders adducted, elbows at 90°, and forearms in a neutral position, following standard procedures established in previous protocols (1, 2). They were instructed to squeeze the device with maximum effort, holding each contraction 3-5 seconds, with 30-second rest intervals between attempts.

**Knee extension strength** of dominant and non-dominant sides was assessed using the VALD DynaMo PLUS handheld dynamometer (VALD Performance, Brisbane, Queensland, Australia). Participants were seated on a comfortable, heavy-duty chair with standardised testing positions in accordance with previously established protocols (3, 4). Each participants performed three maximum effort knee extension per leg, with 30 seconds rest between trials. The average value of three trial from the dominant side (arm and leg) was used in the analysis.

**Gait speed** was assessed using the **10-Meter Walk Test**. Participants were instructed to walk a 10-metre straight path at their usual walking pace. Timing began when the lead foot crossed the start line and ended when it crossed the 10-metre mark. The test was conducted in a level indoor corridor using a stopwatch, and walking aids were permitted if normally used. The time in seconds was recorded to calculate walking speed in metres per second.

**3-m timed-up-and-go testing (TUG) test**. Participants started seated in a standard-height chair. Upon instruction, they stood up, walked 3 metres at a comfortable pace, turned, walked back to the chair, and sat down. The total time to complete the task was recorded in seconds using a stopwatch. The test was performed in a quiet, unobstructed space under the supervision of a trained assessor.

**6-minute walking distance (6MWD),** participants were asked to walk back and forth along a flat, clearly marked 30-metre indoor track for six minutes, covering as much distance as possible. The test was self-paced; resting was allowed, if necessary, although the timer continued. The total distance walked in metres was recorded at the end of the six-minute period, with all assessments supervised in a controlled environment.

**30-Second Chair Stand Test (30CST),** participants sat on a straight-backed chair without armrests and were instructed to stand up fully without using their arms. The test was administered only if the participant could independently stand at least once. Performance was monitored to assess lower limb strength and functional mobility.

### Near Infrared Spectroscopy (NIRS) Derived Mitochondrial Oxidative Capacity (MCO) Modified Test Protocol

Maximum voluntary contraction (MVC) of the handgrip (dominant side) was measured using a Jamar dynamometer in a standardised position. Participants were seated in a comfortable chair without armrest with the elbow and knees flexed at a 90-degree angle and were instructed to exert maximal handgrip force for 3–5 seconds. Strong verbal encouragement was given to maximise the force level as much as possible. E-Link software for Windows (v.16.110) was used to record hand grip tests. Three trials were completed with a 30-second rest interval between the measurements and the average of three peak values was calculated as the final MVC (Jackson et al., 2020).

Recovery of mVO₂ was assessed using a NIRS system (OxiplexTS, ISS, Champaign, USA), which has one light-emitting diode (LED) detector fibre bundle and eight LEDs functioning at wavelengths of 690 and 830 nm (four LEDs per wavelength). A small probe was used, featuring light source-detector distances ranging from 1.5 to 3 centimetres (cm) for each wavelength, with the cell water concentration assumed to be constant at 70%. Data were sampled at a frequency of 2 Hz. NIRS optodes were placed on the bulk of the FDS muscle of the dominant arm (Sumner *et al.*, 2020b; DePauw *et al.*, 2021). To locate the FDS muscle, participants were instructed to resist a pulling force applied to the first four fingers while the contracting muscle was palpated (Vigouroux *et al.*, 2006). Before optode placement, the probe placement area was shaved and cleaned with 70% alcohol wipes to reduce unwanted light reflection. Adipose tissue thickness (ATT) was measured at the site of the NIRS optode using a Harpenden skinfold calliper (British Indicators, Baty International UK). The NIRS device was covered with a soft black cloth to avoid infiltration of ambient light and movement artefacts. According to the manufacturer’s recommendations, the NIRS probe was calibrated before each testing session using a calibration block of known absorption and scattering coefficients. Calibration was then cross-checked using a second block of known but distinctly different absorption and scattering coefficients.

A 10-cm wide blood pressure cuff (Hokanson E20 Rapid Cuff Inflator; Bellevue,98005 WA, USA) was placed over the arm, 2cm above the medial epicondyle of the humerus, and connected to a rapid inflation system (Hokanson AG-101 CUFF INFLATOR AIR SOURCE, Bellevue, WA, USA). The cuff pressures were maintained at 200 mmHg for all occlusion procedures (Gerovasili *et al.*, 2010; Sumner *et al.*, 2020b). A 3-minute arterial occlusion was applied to deoxygenate the tissue beneath the optode to normalise the NIRS signal and record the reperfusion rate. Participants were then asked to rest for 4 minutes to allow the NIRS signal to stabilise and return to baseline values. After a 4-minute recovery period, participants performed a 1-minute isometric handgrip exercise at 50% of their MVC using the same dynamometer setup described earlier. The participants were instructed to perform the test with real-time visual feedback provided by the E-Link software (v. 16.110). Immediately following the exercise, participants released the handgrip and returned their arms to a neutral position alongside the torso within approximately 2-3 seconds, followed by the application of 20 consecutive occlusions (10s for occlusions 1–10 with 10s rest between, 15s for occlusions 11–20 with 15s rest between) (Ryan et al., 2012; Fennell et al., 2023). Data was collected using data acquisition software (OxiTS software version 3.1). It was displayed in real-time and stored for later analysis.

### Gas Chromatography- Mass Spectrometry protocol

**Plasma Sample processing**

Plasma samples were deproteinized in a single batch by adding 300 μl ice cold acetonitrile/methanol (1:1) to 100 μl homogenised plasma spiked with deuterated internal standards, vortexed for 15 seconds and centrifuged at 30130 *g* for 15 minutes. Supernatant was transferred into microcentrifuge tubes (Eppendorf, Stevenage, UK) and lyophilised for 16 h in a vacuum centrifuge (Savant SpeedVac SPD130DLX, Savant vapor trap RVT5105, (Fisher Scientific, Loughborough, UK). Pooled quality control (QC) and process blanks were prepared in tandem.

**Plasma metabolite analysis details**

**Materials**

Glycine-*d*_5_, lysine-*d*_4_, succinic acid-*d*_4_ and alanine-*d*_7_ (98% purity) were obtained from Cambridge Isotopes (Tewksbury, MA) and used as internal standards. All other chemicals were obtained from Fisher Scientific (Loughborough, UK), including LC-MS grade methanol and acetonitrile, pyridine (99% purity), hexane (95% purity), *N*-Methyl-*N*-(trimethylsilyl)trifluoroacetamide and *O*-methylhydroxylamine hydrochloride (98% purity) derivatisation agents, decane, dodecane, pentadecane, nonadecane and docosane (99% purity) retention index alkanes.

**Sample extraction**

Samples were deproteinized in a single batch by adding 300 μl ice cold acetonitrile/methanol (1:1) to 100 μl plasma, spiked with 100 μl of an internal standard mix (1.67 mg/mL of each glycine-*d*_5_, lysine-*d*_4_, succinic acid-*d*_4_ and alanine-*d*_7_ in water) before vortex mixing for 30 s, then centrifuged at 30 130 *g* for 15 min. Supernatant was transferred into microcentrifuge tubes (Eppendorf, Stevenage, UK) and a 15 μl aliquot of each sample was taken to create a pooled quality control (QC). The pooled QC was vortexed for 15 s and aliquoted into microcentrifuge tubes to match sample volume. Process blanks were subjected to the same procedure above. Samples, QCs and process blanks were briefly spun in a centrifuge to remove liquid from vial lids and lyophilised for 16 h in a vacuum centrifuge (Savant SpeedVac SPD130DLX, Savant vapor trap RVT5105, (Fisher Scientific, Loughborough, UK).

**Sample derivatisation**

Samples were divided into randomised batches for derivatisation. Dried extracts were reconstituted in 50 μL of a 20 mg/mL *O*-methylhydroxylamine in pyridine solution, vortex mixed for 10 s, and placed into a digital heat block (Eppendorf, Stevenage, UK) for 40 min, 65 °C. 50 μL *N*-Methyl-*N*-(trimethylsilyl)trifluoroacetamide was then added to each sample, vortex mixed for 10 s, and placed into a digital heat block for 40 min, 65 °C. QC extracts and process blanks were derivatised simultaneously. Derivatised samples were transferred to high recovery GC-MS vials (Agilent, Cheadle, UK) for analysis.

**GC-ToF-MS analysis**

Procedures for GC-MS analysis are adapted from the method presented in (Dunn *et al.*, 2011). A solution containing 0.3 mg/mL of each decane, dodecane, pentadecane, nonadecane and docosane in anhydrous pyridine was prepared and 100 μL added to a high recovery GC-MS vial for us as retention index markers. Retention index solution and process blanks were injected prior to and following the sample run.

QC plasma was injected for capillary column conditioning, followed by further QC injections interspersed with samples, as according to the sequence in Appendix 1. 1 μL of derivatised sample was injected via autosampler through a split/splitless inlet held at 280 °C with a 50:1 split ratio. Autosampler parameters were set to avoid contamination through sequential needle solvent washes and avoid bubble entrainment. A constant helium carrier gas flow of 1 mL/min was maintained with electronic pressure control through a HP-5ms (30 m × 0.25 mm × 0.25 µm) capillary column (Agilent, Cheadle, UK). An oven temperature gradient of 70 °C, 4 min hold, 20 °C/min to 300 °C, 4 min hold was programmed, with a 300 °C transfer line temperature. The mass spectrometer was operated at 70 eV ionisation energy, 200 °C source temperature, 40-600 m/z acquisition range at 10 Hz.

**Data processing**

Raw vendor format data was converted to mzML open format in MSConvert, with a peak picking filter applied to extract centroid data. Files were then imported into MS-DIAL for deconvolution, alignment and annotation. Peak detection was carried out in accurate MS mode, with masses associated with alkanes and trimethylsilyl moieties added to an exclusion list. A retention index dictionary was created by manually integrating peaks from the injected alkane ladder in MassHunter Qualitative Analysis and inputting the associated retention time for each compound into an index file for upload into MS-DIAL. For peak annotation, a retention index tolerance of 20 and *m/z* tolerance of 0.75 Da was utilised, with an EI similarity score cut-off of 70%. An internal electron ionisation (EI) spectral library was used for peak annotation, concatenated with purchased NIST, Fiehn and GOLM libraries. Following deconvolution, alignment and peak annotation, each peak spot was manually inspected for quality by reviewing integrated peak shape, singal:noise ratios, extracted ion chromatograms, raw and deconvolved peak spectra and alignment results. Peak annotations were curated for accuracy by inspecting fragmentation patterns, closest library hits and performing a manual check of retention index similarity and EI similarity scores. Annotation hits from the internal CMR spectral library were preferentially selected, where applicable. Labelled internal standards were utilised in quality checks by examining deconvolved peak area and retention index across samples. Data were exported from MS-DIAL into a tabular matrix containing peak annotations, retention time, assigned retention index and peak area. Signal correction across batches was carried out with QC based robust LOESS (locally estimated scatterplot smoothing) (QC-RLSC). Peaks were filtered by applying a blank sample 10% peak area threshold, D-ratio <40% and QC peak RSD <30%. The resulting batch corrected and filtered peak matrix was used in statistical analysis.

| Injection order | Sample ID |
| --- | --- |
| 1 | Retention index |
| 2 | Blank |
| 3 | Conditioning |
| 4 | Conditioning |
| 5 | Conditioning |
| 6 | Conditioning |
| 7 | Conditioning |
| 8 | QC |
| 9 | QC |
| 10 | Sample |
| 11 | Sample |
| 12 | Sample |
| 13 | Sample |
| 14 | Sample |
| 15 | Sample |
| 16 | Sample |
| 17 | QC |
| 18 | Sample |
| 19 | Sample |
| 20 | Sample |
| 21 | Sample |
| 22 | Sample |
| 23 | Sample |
| 24 | Sample |
| 25 | QC |
| 26 | Sample… |
|  | …continued |
|  | QC |
|  | QC |
|  | Blank |
|  | Retention index |

**Table 1.** GC-MS Data analysis and quality check

| **Software** | | **Version** | **Usage** |
| --- | --- | --- | --- |
| MassHunter Workstation  (Agilent) | | 10.0 | GC-MS instrument control, autosampler control, data acquisition |
| MassHunter Qualitative Analysis (Agilent) | | 10.0 | Manual data quality checks |
| MassHunter Unknowns Analysis (Agilent) | | 10.0 | Spectral deconvolution, EI spectra export to custom library |
| ProteoWizard MSConvert | | 3.0.22286-7583e1c | MS raw file conversion |
| MS-DIAL | | 4.9221218 | Spectral deconvolution, retention index assignment, peak alignment, library matching, signal correction, blank subtraction |
| **Library** | **Version** | | |
| NIST | 20 | | |
| Fiehn | 2013 | | |
| CMR internal | 2024 | | |
| GOLM |  | | |

**Biomarker Assay Protocols**

Six plasma biomarkers were quantified: tumour-necrosis-factor-α (TNF-α) [Thermo Fisher Scientific, Cat. No. KAC1751], interleukin-6 (IL-6) [Thermo Fisher Scientific, Cat. No. EH2IL6], C-reactive protein (CRP) [Thermo Fisher Scientific, Cat. No. KHA0031], glucose [Thermo Fisher Scientific, Cat. No. EIAGLUC], iso-insulin [Thermo Fisher Scientific, Cat. No. BMS2003] and N-terminal pro-B-type natriuretic peptide (NT-proBNP) [Elabscience, Cat. No. E-EL-H6126]. Glucose concentration was determined using a colorimetric detection kit, whereas the remaining biomarkers were quantified by enzyme-linked immunosorbent assay (ELISA) in accordance with the manufacturers’ instructions with small adjustments. All assays were performed in duplicate using appropriate standard curves and controls, with adherence to quality control criteria.

TNF-α: A six-point standard curve (Limit of Detection (LoD) stated to be 0.7 pg/mLpg/mL) was prepared. Samples, standards, and blanks were incubated in duplicate. Following binding, anti-TNF-α HRP conjugate and TMB substrate were applied sequentially. Absorbance was read at 450 nm with background absorbance subtracted. Concentrations were calculated using a 4-parameter logistic (4-PL) regression model.

IL-6: A six-point standard curve (Assay range is stated to be 10.24-400 pg/mL) was generated using serial dilutions. Biotinylated antibody and Streptavidin-HRP were used for detection. Plates were read at 450nm with absorbance at 540 nm subtracted to correct for optical imperfections, and concentrations interpolated using 4-PL regression.

CRP: An eight-point standard curve (Sensitivity is stated as down to <10 pg/mL) was used. Plasma samples were diluted 1:3,000. Assay steps included Biotin Conjugate, Streptavidin-HRP, and Stabilised Chromogen substrate. Readings at 450 nm were analysed via 4-PL regression with backgroumd absorbance subtracted and dilution correction applied. Samples exceeding the detection range were re-assayed at 1:6,000 dilution.

Glucose: Plasma was diluted 1:15. A standard curve (Sensitivity stated down to 0.413 mg/dL) was prepared. After HRP and Glucose Oxidase incubation, absorbance was read at 560 nm with background absorbance subtracted. Final concentrations were calculated via 4-PL regression and adjusted for dilution.

Iso-Insulin: An eight-point standard curve (Sensitivity stated as 6.15 pg/mL) was prepared via 1:2 serial dilutions. Samples and standards were incubated with HRP-conjugate, followed by TMB substrate. Absorbance was read at 450 nm with background absorbance subtracted. Concentrations were calculated via 5-PL regression and corrected for 1:2 dilution.

NT-proBNP: A seven-point standard curve (Detection range stated to be 0.16-10 ng/mL) was prepared by 1:2 serial dilutions. Sequential incubation with biotinylated detection antibody and HRP conjugate was followed by substrate reaction. Plates were read at 450 nm with background absorbance subtracted, and concentrations interpolated using 4-PL regression.

All absorbance measurements were taken using a Multiskan FC microplate reader (Thermo Scientific), and raw data were captured using SkanIt software (v7.1, Drug Discovery Edition). Washing steps were all performed using the Wellwash (Thermo Scientific) and kit-supplied wash solutions. Quality control criteria included R² ≥ 0.99 for standard curves and ±15% deviation for back-calculated values. Samples with a coefficient of variation >20% or exceeding detection limits were re-analysed where further sample was available.

| **Table 2.** Available results for the plasma analysis | | | | |
| --- | --- | --- | --- | --- |
|  | NonHF-Non Frail | NonHF-Frail | HF-NonFrail | HF-Frail |
| GDF-15 | 18 | 6 | 7 | 13^a^ |
| Glucose | 18 | 7 | 7 | 17 |
| Iso-Insulin | 18 | 7 | 7 | 17 |
| NT-ProBNP | 12 | 7 | 5 | 13 |
| TNF-a | 18 | 7 | 7 | 16 |
| Activin A | 15 | 5 | 7 | 3 |
| Follistatin-like 3 | 17 | 5 | 7 | 14 |
| GC-MS | 20 | 6 | 8 | 16 |
| ^a^ results for one participant was excluded to be above detection rate as outlier | | | | |

4. VALD Performance. DynaMo Plus Test: Knee Strength Protocol: VALD; n.d. [Available from: <https://support.vald.com/hc/en-au/articles/19112858914329-DynaMo-Plus-Test-Knee-Strength-Protocol>.

5. IPAQ Research Committee. Scoring protocol for the International Physical Activity Questionnaire (IPAQ) 2005 [Available from: <https://sites.google.com/view/ipaq/score>.

**Table S1.** Pairwise comparison for total HF and total NonHF.

| Metabolites Names | t.stat | p.value | FDR |
| --- | --- | --- | --- |
| 2-Piperidinecarboxylic acid | -0.13222 | 0.89535 | 0.91594 |
| 3-(1-pyrazolyl)-L-alanine | 0.6112 | 0.54389 | 0.66129 |
| 3-Methyl-2-oxopentanoic-acid_1 | -1.7851 | 0.08043 | 0.16343 |
| 4-androsten-3,17-dione 2 | 2.8604 | 0.0062028 | 0.020446 |
| Acetic acid. 4-hydroxyphenyl- | -2.5427 | 0.014209 | 0.040381 |
| Adipic acid. 2-amino- | -0.9175 | 0.36337 | 0.49754 |
| Alanine | -6.4227 | 5.20E-08 | 1.05E-06 |
| Arabinoheptulosonic acid enol. 3-deoxy- BP | -1.6061 | 0.11467 | 0.21808 |
| Arabitol | -0.91393 | 0.36523 | 0.49779 |
| Asparagine | 5.2377 | 3.41E-06 | 3.30E-05 |
| Aspartic acid | 2.4139 | 0.019563 | 0.05213 |
| Benzoic acid. | 0.76915 | 0.4455 | 0.57463 |
| Benzylalcohol | -1.4454 | 0.1547 | 0.26997 |
| Beta-D-Allose | 0.88558 | 0.38017 | 0.51265 |
| Butanoic acid. 2-amino- | 1.9287 | 0.059573 | 0.12869 |
| Butanoic acid. 2-hydroxy- | -1.4717 | 0.14751 | 0.26362 |
| Butanoic acid. 2.4-dihydroxy- | -2.7079 | 0.0092971 | 0.027954 |
| Butanoic acid. 3-hydroxy- | -1.8894 | 0.064759 | 0.13855 |
| Caffeine | 0.65122 | 0.51795 | 0.63495 |
| Carbodiimide | 0.35244 | 0.72602 | 0.79381 |
| Catechol | 0.81747 | 0.41762 | 0.5482 |
| Citric acid | -1.245 | 0.21906 | 0.34691 |
| Creatinine | 4.026 | 0.0001965 | 0.0012318 |
| Cysteine | 4.2244 | 0.0001038 | 0.0006996 |
| Cysteine. S-methyl-. DL- | 0.54053 | 0.59128 | 0.70165 |
| Cysteinyl-glycine | 1.8768 | 0.066506 | 0.1416 |
| Cystine | -3.239 | 0.0021561 | 0.0085668 |
| Cystineamine | -8.1991 | 9.49E-11 | 6.04E-09 |
| Decanoic acid | 0.60526 | 0.5478 | 0.66236 |
| Diethylenglycol | -4.086 | 0.0001622 | 0.0010462 |
| dihydrotestosterone | 0.75811 | 0.45202 | 0.57968 |
| DL- Glutamine | 5.0899 | 5.68E-06 | 5.06E-05 |
| Dodecanoic acid | -1.3985 | 0.16826 | 0.28578 |
| Ergothioneine | -2.776 | 0.0077721 | 0.024356 |
| Erythritol | -2.0537 | 0.045362 | 0.10299 |
| Erythrose | -0.27924 | 0.78123 | 0.83771 |
| Fructose | 3.4832 | 0.001053 | 0.0047815 |
| Fucose | -2.9401 | 0.0049955 | 0.017049 |
| Fumaric acid | -1.2251 | 0.22639 | 0.35724 |
| Furan-2-carboxylic acid | 2.5417 | 0.014247 | 0.040381 |
| Galactonic acid | -1.4835 | 0.14436 | 0.26074 |
| Galactonic acid-1.4-lactone | 9.6191 | 7.19E-13 | 1.07E-10 |
| Galactose | -3.451 | 0.0011591 | 0.0051071 |
| Galactose. 2-amino-2-deoxy | -1.1442 | 0.2581 | 0.39199 |
| Galactose. 2-deoxy | -3.775 | 0.0004329 | 0.002408 |
| Galacturonic acid | -4.0496 | 0.0001823 | 0.0011587 |
| Galacturonic acid-1-phosphate | -2.7115 | 0.0092113 | 0.027885 |
| Glucoheptonic acid-1.4-lactone | -1.0228 | 0.31142 | 0.4514 |
| Gluconic acid-1.4-lactone | 4.7595 | 1.76E-05 | 0.0001421 |
| Glucopyranose | -1.2771 | 0.20759 | 0.33335 |
| Glucose. 2-deoxy | -2.5695 | 0.013277 | 0.038313 |
| Glutamic acid | -3.9178 | 0.0002769 | 0.0016215 |
| Glutamine | 0.3157 | 0.75357 | 0.8179 |
| Glutaric acid | -2.0586 | 0.044868 | 0.10239 |
| Glutaric acid. 2-hydroxy- | -0.99214 | 0.32601 | 0.46163 |
| Glutaric acid. 2-oxo- | -3.6554 | 0.0006257 | 0.0032757 |
| Glyceric acid | -1.3353 | 0.18794 | 0.3144 |
| Glycerol | -4.1361 | 0.0001381 | 0.0009036 |
| Glycine | 1.5824 | 0.12 | 0.22531 |
| Glycine.1 | 2.4161 | 0.01946 | 0.05213 |
| Glycolic acid | 0.16496 | 0.86965 | 0.89999 |
| Glyoxylic acid | 4.2516 | 9.50E-05 | 0.0006502 |
| Heptadecanoic acid | 0.56351 | 0.57566 | 0.68678 |
| Hippuric acid | -1.7798 | 0.081311 | 0.16447 |
| Histidine | 6.244 | 9.83E-08 | 1.82E-06 |
| Histidine. N-tau-methyl- | 1.819 | 0.07503 | 0.15498 |
| Hydroquinone | 2.1961 | 0.032846 | 0.080311 |
| Hypotaurine | -1.0956 | 0.27861 | 0.4119 |
| Hypoxanthine | -1.1068 | 0.27377 | 0.40609 |
| Idose | -6.1408 | 1.42E-07 | 2.49E-06 |
| Indole-3-acetamide | 2.0592 | 0.044808 | 0.10239 |
| Indole-3-acetic acid | -0.98554 | 0.3292 | 0.46359 |
| Indole. 3-hydroxy- | -1.6126 | 0.11325 | 0.2163 |
| Isobutanoic acid. 3-amino- | -2.1769 | 0.034335 | 0.082589 |
| Isocaproic acid. 2-oxo | -1.1935 | 0.23842 | 0.36967 |
| Isoleucine | -3.6082 | 0.0007224 | 0.0035719 |
| Isomaltose | -3.2405 | 0.0021472 | 0.0085668 |
| isopropyl beta-D-1-thiogalactopyranoside [19.097] | -1.4481 | 0.15396 | 0.26973 |
| Isovaleric acid. 2-oxo | -2.4221 | 0.019174 | 0.051711 |
| Lactic acid | 1.4735 | 0.147 | 0.26362 |
| Lactic acid. 3-(4-hydroxyphenyl)- | -0.71011 | 0.481 | 0.60126 |
| Loganin | -1.1797 | 0.24382 | 0.37543 |
| Lysine | -1.3564 | 0.18118 | 0.30425 |
| Malic acid | -3.3366 | 0.0016244 | 0.0067558 |
| Malonic acid | -2.3433 | 0.023214 | 0.059812 |
| Mannopyranoside | -0.1047 | 0.91704 | 0.93169 |
| Methionine | -4.7151 | 2.04E-05 | 0.0001565 |
| Methoxytryptamine | -1.6762 | 0.10007 | 0.19447 |
| Muramic acid. N-acetyl | 3.3849 | 0.0014096 | 0.0060443 |
| Myo-Inositol | 2.9778 | 0.0045037 | 0.015781 |
| N-acetyl Galactosamine | -4.0146 | 0.0002038 | 0.0012594 |
| N-acetyl- Aspartic acid | -6.1343 | 1.45E-07 | 2.49E-06 |
| N-Carboxyglycine | 5.0894 | 5.69E-06 | 5.06E-05 |
| N-methyl trans-4-hydroxy-L-proline (2S.4R)-4-hydroxy-1-methyl pyrrolidine-2-carboxylic acid | 2.0743 | 0.043327 | 0.099899 |
| Neryl acetate | -0.36044 | 0.72007 | 0.79119 |
| Neuraminic acid. N-glycolyl | -0.21165 | 0.83326 | 0.87453 |
| Nivalenol | 3.8609 | 0.0003312 | 0.0018655 |
| Nonanoic acid | 0.36062 | 0.71993 | 0.79119 |
| Norleucine | 2.5307 | 0.014645 | 0.041246 |
| Norvaline | 2.0833 | 0.042462 | 0.098415 |
| O-Toluic-acid | 0.49413 | 0.62343 | 0.72246 |
| Octadecadienoic acid | 3.7536 | 0.0004626 | 0.0025411 |
| Octadecenoic acid | 2.1397 | 0.037387 | 0.088969 |
| Octadecenoic acid methyl ester | 5.8686 | 3.73E-07 | 5.18E-06 |
| Octanoic acid | 0.99055 | 0.32677 | 0.46163 |
| Ornithine | -3.5228 | 0.0009352 | 0.0044275 |
| Oxalic acid | 0.22816 | 0.82047 | 0.86774 |
| Pantothenic acid | 0.82624 | 0.41267 | 0.54386 |
| Pentanoic acid. 2-propy | -0.43394 | 0.66623 | 0.75409 |
| Phenol | 0.15102 | 0.88058 | 0.90919 |
| Phenylalanine | -5.4738 | 1.50E-06 | 1.75E-05 |
| Phosphoric acid | 0.72017 | 0.47484 | 0.5969 |
| Phosphoric acid monomethyl ester | 3.4603 | 0.0011273 | 0.0050164 |
| Proline | -0.62299 | 0.53618 | 0.6555 |
| Proline. 4-hydroxy | 2.6311 | 0.011346 | 0.033886 |
| Propane-1.2-diol | -1.8532 | 0.069883 | 0.14718 |
| Psicose | -1.1728 | 0.24655 | 0.37772 |
| Pyridine. 2-hydroxy- | 2.62 | 0.011674 | 0.034634 |
| Pyridine. 3-hydroxy- | 3.2295 | 0.002216 | 0.0087268 |
| Pyroglutamic acid | 0.25627 | 0.79881 | 0.85245 |
| Pyruvic acid | -1.7491 | 0.08653 | 0.17114 |
| Ribitol | -0.70561 | 0.48377 | 0.60289 |
| Ribonic acid | -1.7721 | 0.0826 | 0.1657 |
| Ribose | 0.76957 | 0.44525 | 0.57463 |
| Salicin | -1.6147 | 0.11279 | 0.2163 |
| Salicylic acid | -1.5542 | 0.12658 | 0.23666 |
| Sarcosine | 1.599 | 0.11625 | 0.22013 |
| Sarcosineamide | 1.0417 | 0.30268 | 0.44161 |
| Sedoheptulose. 2.7-anhydro-. beta- | 3.5166 | 0.0009526 | 0.0044443 |
| Serine | -1.016 | 0.31463 | 0.45348 |
| Serine. O-phospho- | 0.53243 | 0.59683 | 0.70494 |
| Siloxane | 1.3056 | 0.19779 | 0.32333 |
| similar to Asparagine Derivate | 5.579 | 1.03E-06 | 1.24E-05 |
| similar to Glycerolaldopyranosid | -1.4291 | 0.15932 | 0.276 |
| similar to Pentasiloxane. dodecamethyl | -6.6557 | 2.27E-08 | 6.04E-07 |
| Sorbose | -0.308 | 0.75939 | 0.82221 |
| Tartaric acid | 2.4634 | 0.017321 | 0.047579 |
| Tartronic acid | -0.97936 | 0.33221 | 0.46635 |
| Tetradecanoic acid | -1.5413 | 0.12968 | 0.24145 |
| Threonic acid-1.4-lactone | 2.8138 | 0.0070277 | 0.022662 |
| Threonine | 2.6123 | 0.011906 | 0.035086 |
| Tryptophan | 3.2624 | 0.0020156 | 0.008229 |
| Tyrosine | -1.932 | 0.059159 | 0.12842 |
| Tyrosine.1 | 1.6771 | 0.099896 | 0.19447 |
| Unknown01 | -0.46443 | 0.6444 | 0.73716 |
| Unknown02 | 3.2058 | 0.0023723 | 0.0091796 |
| Unknown03 | 2.0955 | 0.041317 | 0.09677 |
| Unknown04 | 2.2516 | 0.028863 | 0.071755 |
| Unknown05 | 3.7232 | 0.000508 | 0.002757 |
| Unknown06 | 1.7504 | 0.086313 | 0.17114 |
| Unknown07 | 2.5978 | 0.012357 | 0.036177 |
| Unknown08 | -5.1502 | 4.61E-06 | 4.28E-05 |
| Unknown09 | 2.8835 | 0.005827 | 0.019496 |
| Unknown10 | -1.4288 | 0.1594 | 0.276 |
| Unknown100 | 1.2168 | 0.2295 | 0.36087 |
| Unknown101 | -1.305 | 0.19797 | 0.32333 |
| Unknown102 | 1.1946 | 0.238 | 0.36967 |
| Unknown103 | -0.02459 | 0.98048 | 0.98048 |
| Unknown104 | -3.879 | 0.0003129 | 0.0018081 |
| Unknown105 | -1.2596 | 0.21377 | 0.33974 |
| Unknown106 | 4.3269 | 7.43E-05 | 0.0005163 |
| Unknown107 | 0.13256 | 0.89509 | 0.91594 |
| Unknown108 | 11.063 | 6.34E-15 | 1.41E-12 |
| Unknown109 | 3.1156 | 0.0030663 | 0.011564 |
| Unknown11 | 0.23002 | 0.81903 | 0.86774 |
| Unknown110 | -5.7164 | 6.38E-07 | 8.22E-06 |
| Unknown111 | -5.9476 | 2.82E-07 | 4.18E-06 |
| Unknown112 | -4.9683 | 8.62E-06 | 7.38E-05 |
| Unknown113 | 0.52547 | 0.60162 | 0.70648 |
| Unknown114 | 1.8177 | 0.075227 | 0.15498 |
| Unknown115 | -5.5799 | 1.03E-06 | 1.24E-05 |
| Unknown116 | -1.7867 | 0.080179 | 0.16343 |
| Unknown117 | -2.4251 | 0.019037 | 0.051655 |
| Unknown118 | 0.47536 | 0.63664 | 0.73395 |
| Unknown119 | 2.0846 | 0.042339 | 0.098415 |
| Unknown12 | 6.5705 | 3.07E-08 | 6.83E-07 |
| Unknown120 | 2.786 | 0.0075686 | 0.023887 |
| Unknown121 | 0.93783 | 0.35293 | 0.48624 |
| Unknown122 | -1.3034 | 0.19853 | 0.32333 |
| Unknown123 | -4.4262 | 5.35E-05 | 0.0003906 |
| Unknown124 | -5.353 | 2.28E-06 | 2.42E-05 |
| Unknown125 | 2.2898 | 0.026375 | 0.06631 |
| Unknown126 | -0.77349 | 0.44295 | 0.57463 |
| Unknown127 | -2.4727 | 0.016926 | 0.046782 |
| Unknown128 | 0.24141 | 0.81025 | 0.86258 |
| Unknown129 | -0.29043 | 0.77271 | 0.83156 |
| Unknown13 | 6.3835 | 5.98E-08 | 1.16E-06 |
| Unknown130 | -0.69044 | 0.49318 | 0.61132 |
| Unknown131 | -1.0154 | 0.31489 | 0.45348 |
| Unknown132 | 1.5345 | 0.13134 | 0.24252 |
| Unknown133 | 2.3403 | 0.023387 | 0.059812 |
| Unknown134 | 2.0287 | 0.047936 | 0.10828 |
| Unknown135 | -6.8853 | 9.99E-09 | 3.17E-07 |
| Unknown136 | 5.029 | 7.00E-06 | 6.11E-05 |
| Unknown137 | 2.3449 | 0.023126 | 0.059812 |
| Unknown138 | 0.067417 | 0.94652 | 0.95946 |
| Unknown139 | 0.58763 | 0.55948 | 0.66927 |
| Unknown14 | 1.9589 | 0.055826 | 0.12178 |
| Unknown140 | 6.0674 | 1.84E-07 | 2.93E-06 |
| Unknown141 | 0.83607 | 0.40717 | 0.54249 |
| Unknown142 | 0.91834 | 0.36294 | 0.49754 |
| Unknown143 | -0.68131 | 0.49888 | 0.61667 |
| Unknown144 | -1.3257 | 0.19107 | 0.31608 |
| Unknown145 | 0.83011 | 0.4105 | 0.54367 |
| Unknown146 | -1.9744 | 0.053987 | 0.11952 |
| Unknown147 | 1.1978 | 0.23677 | 0.36967 |
| Unknown148 | 0.027913 | 0.97784 | 0.98005 |
| Unknown149 | 2.8227 | 0.0068636 | 0.022294 |
| Unknown15 | 5.1649 | 4.38E-06 | 4.15E-05 |
| Unknown150 | 0.50591 | 0.61519 | 0.71478 |
| Unknown151 | -2.3254 | 0.024234 | 0.061273 |
| Unknown152 | -0.38725 | 0.70025 | 0.77712 |
| Unknown153 | -0.94837 | 0.3476 | 0.48037 |
| Unknown154 | -0.14741 | 0.88341 | 0.91 |
| Unknown155 | -1.3086 | 0.19677 | 0.32333 |
| Unknown156 | 0.22601 | 0.82213 | 0.86774 |
| Unknown157 | -2.2229 | 0.030869 | 0.076314 |
| Unknown158 | 0.051787 | 0.95891 | 0.9672 |
| Unknown159 | -1.5104 | 0.13737 | 0.25052 |
| Unknown16 | -0.33803 | 0.73678 | 0.80163 |
| Unknown160 | -1.6188 | 0.11191 | 0.21559 |
| Unknown161 | -0.43033 | 0.66884 | 0.75409 |
| Unknown162 | -6.03 | 2.10E-07 | 3.23E-06 |
| Unknown163 | -1.9715 | 0.054324 | 0.11967 |
| Unknown164 | 1.1818 | 0.243 | 0.37543 |
| Unknown165 | -2.508 | 0.015504 | 0.043216 |
| Unknown166 | -0.83269 | 0.40906 | 0.54338 |
| Unknown167 | -5.4381 | 1.70E-06 | 1.93E-05 |
| Unknown168 | -1.1265 | 0.26544 | 0.40003 |
| Unknown169 | 0.19056 | 0.84965 | 0.88547 |
| Unknown17 | 3.3374 | 0.0016203 | 0.0067558 |
| Unknown170 | -36.169 | 5.28E-37 | 2.35E-34 |
| Unknown171 | -1.0101 | 0.3174 | 0.45416 |
| Unknown172 | 1.4868 | 0.14348 | 0.2606 |
| Unknown173 | 1.8312 | 0.073152 | 0.15212 |
| Unknown174 | 1.8549 | 0.069633 | 0.14718 |
| Unknown175 | 1.9949 | 0.051626 | 0.11603 |
| Unknown176 | -0.74986 | 0.45692 | 0.58428 |
| Unknown177 | 2.8365 | 0.0066143 | 0.021642 |
| Unknown178 | 2.7969 | 0.0073529 | 0.02354 |
| Unknown179 | -0.67088 | 0.50545 | 0.62286 |
| Unknown18 | -5.8688 | 3.73E-07 | 5.18E-06 |
| Unknown180 | -2.0997 | 0.040928 | 0.096364 |
| Unknown181 | 1.111 | 0.27201 | 0.40483 |
| Unknown182 | 0.91007 | 0.36724 | 0.49824 |
| Unknown183 | 1.7703 | 0.082905 | 0.1657 |
| Unknown184 | 1.4082 | 0.16539 | 0.28416 |
| Unknown185 | 3.0227 | 0.0039781 | 0.014162 |
| Unknown186 | 3.9365 | 0.0002611 | 0.0015489 |
| Unknown187 | -8.8465 | 1.00E-11 | 9.89E-10 |
| Unknown188 | -3.0775 | 0.0034129 | 0.012449 |
| Unknown189 | -1.8913 | 0.064504 | 0.13855 |
| Unknown19 | 5.402 | 1.92E-06 | 2.14E-05 |
| Unknown190 | -4.5492 | 3.56E-05 | 0.000264 |
| Unknown191 | 2.1143 | 0.039599 | 0.093731 |
| Unknown192 | -4.3891 | 6.05E-05 | 0.0004275 |
| Unknown193 | 2.5675 | 0.013345 | 0.038313 |
| Unknown194 | 1.2956 | 0.20117 | 0.32553 |
| Unknown195 | -6.5957 | 2.81E-08 | 6.58E-07 |
| Unknown196 | 0.8255 | 0.41309 | 0.54386 |
| Unknown197 | -0.77905 | 0.4397 | 0.57212 |
| Unknown198 | 0.47036 | 0.64019 | 0.73424 |
| Unknown199 | -0.66891 | 0.50669 | 0.62286 |
| Unknown20 | -0.59711 | 0.55319 | 0.66532 |
| Unknown200 | -7.764 | 4.39E-10 | 2.44E-08 |
| Unknown201 | 0.51637 | 0.60792 | 0.71004 |
| Unknown202 | 3.3381 | 0.0016171 | 0.0067558 |
| Unknown203 | -0.37034 | 0.71273 | 0.78701 |
| Unknown204 | -3.0485 | 0.0037019 | 0.013393 |
| Unknown205 | -1.4694 | 0.14812 | 0.26365 |
| Unknown206 | -1.2752 | 0.20825 | 0.33335 |
| Unknown207 | -0.37762 | 0.70734 | 0.783 |
| Unknown208 | 0.73878 | 0.46356 | 0.58729 |
| Unknown209 | 0.43191 | 0.6677 | 0.75409 |
| Unknown21 | 4.1603 | 0.0001277 | 0.0008481 |
| Unknown210 | 2.1792 | 0.034155 | 0.082589 |
| Unknown211 | -1.3018 | 0.19908 | 0.32333 |
| Unknown212 | 6.7454 | 1.65E-08 | 4.88E-07 |
| Unknown213 | 3.8672 | 0.0003247 | 0.0018527 |
| Unknown214 | -0.049557 | 0.96068 | 0.9672 |
| Unknown215 | 1.6799 | 0.099347 | 0.19447 |
| Unknown216 | 1.3294 | 0.18986 | 0.31525 |
| Unknown217 | -1.519 | 0.13519 | 0.24758 |
| Unknown218 | -0.39821 | 0.69221 | 0.77395 |
| Unknown219 | 0.14456 | 0.88565 | 0.91019 |
| Unknown22 | -1.9907 | 0.052101 | 0.11613 |
| Unknown220 | 4.6398 | 2.63E-05 | 0.0001983 |
| Unknown221 | -3.5145 | 0.0009588 | 0.0044443 |
| Unknown222 | 0.20494 | 0.83847 | 0.87793 |
| Unknown223 | 1.9899 | 0.052193 | 0.11613 |
| Unknown224 | 0.53187 | 0.59722 | 0.70494 |
| Unknown225 | 2.9384 | 0.0050188 | 0.017049 |
| Unknown226 | 0.70359 | 0.48502 | 0.60289 |
| Unknown227 | 0.71367 | 0.47882 | 0.60021 |
| Unknown228 | 3.6691 | 0.0005999 | 0.0031781 |
| Unknown229 | 0.2218 | 0.82539 | 0.86832 |
| Unknown23 | 0.28922 | 0.77363 | 0.83156 |
| Unknown230 | 2.9946 | 0.0042994 | 0.015184 |
| Unknown231 | 0.35555 | 0.72371 | 0.79322 |
| Unknown232 | 1.2799 | 0.20661 | 0.33311 |
| Unknown233 | -0.51115 | 0.61154 | 0.7124 |
| Unknown234 | 0.91285 | 0.36579 | 0.49779 |
| Unknown235 | 0.4312 | 0.66821 | 0.75409 |
| Unknown236 | -0.5253 | 0.60174 | 0.70648 |
| Unknown237 | 1.7104 | 0.093517 | 0.18414 |
| Unknown238 | 2.2686 | 0.027734 | 0.069336 |
| Unknown239 | 0.97479 | 0.33445 | 0.46734 |
| Unknown24 | -1.5373 | 0.13064 | 0.24223 |
| Unknown240 | 0.22503 | 0.82289 | 0.86774 |
| Unknown241 | 0.40531 | 0.68702 | 0.77008 |
| Unknown242 | 0.85035 | 0.39927 | 0.53516 |
| Unknown243 | 0.041534 | 0.96704 | 0.9714 |
| Unknown244 | -1.6652 | 0.10226 | 0.19785 |
| Unknown245 | -0.88833 | 0.37871 | 0.51223 |
| Unknown246 | -3.1043 | 0.0031653 | 0.011738 |
| Unknown247 | -1.402 | 0.16722 | 0.28578 |
| Unknown248 | -4.846 | 1.31E-05 | 0.0001079 |
| Unknown249 | 3.2932 | 0.0018435 | 0.007596 |
| Unknown25 | 1.8053 | 0.077173 | 0.15826 |
| Unknown250 | 0.95703 | 0.34325 | 0.47734 |
| Unknown251 | -3.5007 | 0.0009992 | 0.0045837 |
| Unknown252 | -3.2548 | 0.0020606 | 0.0083361 |
| Unknown253 | 3.6489 | 0.0006383 | 0.0032924 |
| Unknown254 | 5.7126 | 6.47E-07 | 8.22E-06 |
| Unknown255 | 3.0257 | 0.0039447 | 0.014156 |
| Unknown256 | 1.5247 | 0.13376 | 0.24597 |
| Unknown257 | 2.1613 | 0.03559 | 0.085148 |
| Unknown258 | 0.94943 | 0.34706 | 0.48037 |
| Unknown259 | 4.9304 | 9.82E-06 | 8.24E-05 |
| Unknown26 | -1.4149 | 0.16342 | 0.28187 |
| Unknown260 | 1.1331 | 0.26268 | 0.39759 |
| Unknown261 | 4.7412 | 1.87E-05 | 0.0001458 |
| Unknown262 | -0.48936 | 0.62677 | 0.72445 |
| Unknown263 | 4.0014 | 0.0002125 | 0.0012955 |
| Unknown264 | -0.42961 | 0.66936 | 0.75409 |
| Unknown265 | 0.052064 | 0.95869 | 0.9672 |
| Unknown266 | 3.0853 | 0.0033389 | 0.01228 |
| Unknown267 | 0.99656 | 0.32387 | 0.46163 |
| Unknown268 | 2.1921 | 0.03315 | 0.080609 |
| Unknown269 | -1.3776 | 0.17458 | 0.29427 |
| Unknown27 | 1.3818 | 0.17329 | 0.2932 |
| Unknown270 | -1.4628 | 0.14991 | 0.26578 |
| Unknown271 | -1.8516 | 0.070116 | 0.14718 |
| Unknown272 | 0.26458 | 0.79244 | 0.84768 |
| Unknown273 | 3.2122 | 0.002329 | 0.0090914 |
| Unknown274 | -1.4512 | 0.1531 | 0.26928 |
| Unknown275 | 0.29149 | 0.77191 | 0.83156 |
| Unknown276 | 0.17042 | 0.86538 | 0.89766 |
| Unknown277 | -1.0117 | 0.31664 | 0.45416 |
| Unknown278 | -3.4777 | 0.0010704 | 0.0048115 |
| Unknown279 | 2.3302 | 0.023959 | 0.060923 |
| Unknown28 | -4.4028 | 5.78E-05 | 0.0004151 |
| Unknown280 | -2.7626 | 0.0080516 | 0.025056 |
| Unknown281 | -2.7165 | 0.0090914 | 0.027885 |
| Unknown282 | -1.1716 | 0.247 | 0.37772 |
| Unknown283 | -5.2399 | 3.38E-06 | 3.30E-05 |
| Unknown284 | 0.11556 | 0.90848 | 0.92511 |
| Unknown285 | -2.9403 | 0.0049925 | 0.017049 |
| Unknown286 | -3.1515 | 0.00277 | 0.010615 |
| Unknown288 | -1.1208 | 0.26784 | 0.40003 |
| Unknown289 | 0.73752 | 0.46433 | 0.58729 |
| Unknown29 | -0.76647 | 0.44707 | 0.57499 |
| Unknown290 | 6.4466 | 4.78E-08 | 1.01E-06 |
| Unknown291 | -6.1055 | 1.61E-07 | 2.65E-06 |
| Unknown292 | 8.8157 | 1.11E-11 | 9.89E-10 |
| Unknown293 | -2.4524 | 0.0178 | 0.048595 |
| Unknown294 | 1.1948 | 0.23793 | 0.36967 |
| Unknown295 | -2.9344 | 0.0050731 | 0.017103 |
| Unknown296 | 0.41476 | 0.68013 | 0.76429 |
| Unknown30 | 0.38721 | 0.70028 | 0.77712 |
| Unknown31 | -0.99405 | 0.32508 | 0.46163 |
| Unknown32 | 6.6257 | 2.52E-08 | 6.24E-07 |
| Unknown33 | -1.9596 | 0.055746 | 0.12178 |
| Unknown34 | 3.6102 | 0.000718 | 0.0035719 |
| Unknown35 | 3.5986 | 0.0007437 | 0.003637 |
| Unknown36 | 1.3986 | 0.16823 | 0.28578 |
| Unknown37 | -4.7412 | 1.87E-05 | 0.0001458 |
| Unknown38 | 3.5731 | 0.0008036 | 0.0038871 |
| Unknown39 | -1.7695 | 0.083034 | 0.1657 |
| Unknown40 | -2.3507 | 0.022809 | 0.059705 |
| Unknown41 | -2.9678 | 0.00463 | 0.016097 |
| Unknown42 | -3.6455 | 0.0006448 | 0.0032924 |
| Unknown43 | -0.59073 | 0.55742 | 0.6686 |
| Unknown44 | -0.84664 | 0.40131 | 0.53629 |
| Unknown45 | -0.39419 | 0.69515 | 0.7753 |
| Unknown46 | 8.7543 | 1.37E-11 | 1.02E-09 |
| Unknown47 | 0.34426 | 0.73213 | 0.79852 |
| Unknown48 | -0.74071 | 0.4624 | 0.58729 |
| Unknown49 | -1.0289 | 0.30858 | 0.44875 |
| Unknown50 | -1.0479 | 0.29983 | 0.4389 |
| Unknown51 | 0.12582 | 0.90039 | 0.91898 |
| Unknown52 | 0.61177 | 0.54352 | 0.66129 |
| Unknown53 | -2.7341 | 0.0086815 | 0.026828 |
| Unknown54 | 1.0615 | 0.29368 | 0.43131 |
| Unknown55 | 3.5228 | 0.0009352 | 0.0044275 |
| Unknown56 | 7.1366 | 4.08E-09 | 1.65E-07 |
| Unknown57 | 0.60308 | 0.54924 | 0.66236 |
| Unknown58 | -2.7133 | 0.0091683 | 0.027885 |
| Unknown59 | -3.9562 | 0.0002453 | 0.0014754 |
| Unknown60 | -1.8333 | 0.072837 | 0.15212 |
| Unknown61 | -1.4821 | 0.14472 | 0.26074 |
| Unknown62 | -0.78279 | 0.43752 | 0.57096 |
| Unknown63 | 0.52306 | 0.60329 | 0.70648 |
| Unknown64 | -3.1085 | 0.0031279 | 0.011697 |
| Unknown65 | 1.1228 | 0.267 | 0.40003 |
| Unknown66 | 0.4578 | 0.64912 | 0.74066 |
| Unknown67 | -3.1488 | 0.002791 | 0.010615 |
| Unknown68 | 0.6085 | 0.54567 | 0.66164 |
| Unknown69 | -2.2097 | 0.03183 | 0.078257 |
| Unknown70 | -1.1566 | 0.25305 | 0.38564 |
| Unknown71 | 6.6508 | 2.31E-08 | 6.04E-07 |
| Unknown72 | 2.3718 | 0.021672 | 0.057066 |
| Unknown73 | -0.72347 | 0.47283 | 0.59606 |
| Unknown74 | 5.3133 | 2.62E-06 | 2.65E-05 |
| Unknown75 | 1.1207 | 0.26789 | 0.40003 |
| Unknown76 | 1.3298 | 0.18973 | 0.31525 |
| Unknown77 | 2.3945 | 0.020512 | 0.054332 |
| Unknown78 | 0.19771 | 0.84409 | 0.88173 |
| Unknown79 | -2.3418 | 0.023298 | 0.059812 |
| Unknown80 | 0.73714 | 0.46455 | 0.58729 |
| Unknown81 | -3.71 | 0.0005292 | 0.0028372 |
| Unknown82 | -0.97365 | 0.33501 | 0.46734 |
| Unknown83 | -3.3842 | 0.0014126 | 0.0060443 |
| Unknown84 | 5.3602 | 2.23E-06 | 2.42E-05 |
| Unknown85 | 1.0903 | 0.2809 | 0.41392 |
| Unknown86 | 1.587 | 0.11894 | 0.22427 |
| Unknown87 | -0.17819 | 0.8593 | 0.89344 |
| Unknown88 | -0.4704 | 0.64015 | 0.73424 |
| Unknown89 | 6.9199 | 8.83E-09 | 3.15E-07 |
| Unknown90 | 0.85884 | 0.39461 | 0.53052 |
| Unknown91 | 5.3274 | 2.49E-06 | 2.58E-05 |
| Unknown92 | -3.4393 | 0.0011999 | 0.005235 |
| Unknown93 | -7.4601 | 1.29E-09 | 6.38E-08 |
| Unknown94 | 5.7562 | 5.55E-07 | 7.48E-06 |
| Unknown95 | -7.1775 | 3.52E-09 | 1.57E-07 |
| Unknown96 | -6.9086 | 9.19E-09 | 3.15E-07 |
| Unknown97 | 0.55509 | 0.58136 | 0.69172 |
| Unknown98 | 3.6423 | 0.0006511 | 0.0032924 |
| Unknown99 | 2.787 | 0.007549 | 0.023887 |
| Urea | -2.5696 | 0.013274 | 0.038313 |
| Uric acid | 0.81484 | 0.41911 | 0.54854 |
| Valeric acid. 2-oxo- | -2.5071 | 0.015538 | 0.043216 |
| Valine | -2.8753 | 0.0059584 | 0.019787 |
| Xanthine | -1.4591 | 0.15091 | 0.26649 |
| Xylitol | 1.27 | 0.21009 | 0.33509 |

**Table S2.** Pairwise comparison for HF-Frail and NonHF-NonFrail.

|  | t.stat | p.value | FDR |
| --- | --- | --- | --- |
| 2-Piperidinecarboxylic acid | 0.20208 | 0.84102 | 0.90182 |
| 3-(1-pyrazolyl)-L-alanine | 0.452 | 0.65406 | 0.76073 |
| 3-Methyl-2-oxopentanoic-acid_1 | -1.9596 | 0.058049 | 0.12185 |
| 4-androsten-3,17-dione 2 | 3.3986 | 0.0017046 | 0.0073375 |
| Acetic acid. 4-hydroxyphenyl- | -2.3472 | 0.024703 | 0.062815 |
| Adipic acid. 2-amino- | -1.4721 | 0.14994 | 0.25701 |
| Alanine | -6.0319 | 7.01E-07 | 1.51E-05 |
| Arabinoheptulosonic acid enol. 3-deoxy- BP | -1.3547 | 0.18421 | 0.29593 |
| Arabitol | -1.7212 | 0.094053 | 0.1804 |
| Asparagine | 4.1247 | 0.00021732 | 0.0014434 |
| Aspartic acid | 0.78625 | 0.43701 | 0.56697 |
| Benzoic acid. | 1.745 | 0.089756 | 0.17366 |
| Benzylalcohol | -1.74 | 0.090644 | 0.17462 |
| Beta-D-Allose | 0.58262 | 0.56388 | 0.69317 |
| Butanoic acid. 2-amino- | 1.3722 | 0.17874 | 0.29076 |
| Butanoic acid. 2-hydroxy- | -1.2646 | 0.21437 | 0.33239 |
| Butanoic acid. 2.4-dihydroxy- | -3.3854 | 0.0017669 | 0.0074882 |
| Butanoic acid. 3-hydroxy- | -1.1051 | 0.27664 | 0.39712 |
| Caffeine | 0.84016 | 0.40652 | 0.53572 |
| Carbodiimide | -0.61258 | 0.54412 | 0.67259 |
| Catechol | 0.19064 | 0.84991 | 0.90481 |
| Citric acid | -2.7321 | 0.0097962 | 0.030555 |
| Creatinine | 3.3098 | 0.0021713 | 0.0087839 |
| Cysteine | 3.4318 | 0.0015558 | 0.0068549 |
| Cysteine. S-methyl-. DL- | 0.097944 | 0.92254 | 0.9525 |
| Cysteinyl-glycine | 1.7511 | 0.088682 | 0.17309 |
| Cystine | -3.6283 | 0.00090118 | 0.00436 |
| Cystineamine | -7.1566 | 2.40E-08 | 1.19E-06 |
| Decanoic acid | 0.28247 | 0.77925 | 0.8626 |
| Diethylenglycol | -5.0853 | 1.24E-05 | 0.00013136 |
| dihydrotestosterone | 1.1195 | 0.27057 | 0.38966 |
| DL- Glutamine | 4.4445 | 8.48E-05 | 0.00068606 |
| Dodecanoic acid | -1.2417 | 0.22259 | 0.34038 |
| Ergothioneine | -3.3325 | 0.0020412 | 0.0083334 |
| Erythritol | -2.6313 | 0.012564 | 0.036782 |
| Erythrose | 0.42159 | 0.6759 | 0.78124 |
| Fructose | 2.1921 | 0.035117 | 0.082071 |
| Fucose | -3.1015 | 0.0037925 | 0.014182 |
| Fumaric acid | -2.7785 | 0.0087214 | 0.028123 |
| Furan-2-carboxylic acid | 1.4323 | 0.16092 | 0.27023 |
| Galactonic acid | -2.9578 | 0.0055215 | 0.019347 |
| Galactonic acid-1.4-lactone | 8.0723 | 1.67E-09 | 2.11E-07 |
| Galactose | -3.9166 | 0.00039706 | 0.0023249 |
| Galactose. 2-amino-2-deoxy | -1.4357 | 0.15998 | 0.26966 |
| Galactose. 2-deoxy | -4.1633 | 0.00019416 | 0.00135 |
| Galacturonic acid | -4.9885 | 1.66E-05 | 0.00016066 |
| Galacturonic acid-1-phosphate | -1.9242 | 0.062497 | 0.12935 |
| Glucoheptonic acid-1.4-lactone | -1.3722 | 0.17872 | 0.29076 |
| Gluconic acid-1.4-lactone | 3.8295 | 0.00050979 | 0.0029084 |
| Glucopyranose | -1.2448 | 0.22147 | 0.34038 |
| Glucose. 2-deoxy | -2.1212 | 0.041064 | 0.090462 |
| Glutamic acid | -4.1387 | 0.00020859 | 0.0014253 |
| Glutamine | -0.39154 | 0.69777 | 0.79414 |
| Glutaric acid | -2.3896 | 0.022385 | 0.060014 |
| Glutaric acid. 2-hydroxy- | -1.8318 | 0.075505 | 0.14976 |
| Glutaric acid. 2-oxo- | -5.2232 | 8.17E-06 | 9.32E-05 |
| Glyceric acid | -2.1276 | 0.04049 | 0.089642 |
| Glycerol | -3.5781 | 0.0010371 | 0.0049096 |
| Glycine | 1.4863 | 0.14615 | 0.25306 |
| Glycine.1 | 2.5875 | 0.013981 | 0.040138 |
| Glycolic acid | 0.053286 | 0.95781 | 0.96879 |
| Glyoxylic acid | 3.7523 | 0.00063506 | 0.0034048 |
| Heptadecanoic acid | 1.2082 | 0.23509 | 0.35245 |
| Hippuric acid | -2.2909 | 0.028106 | 0.068769 |
| Histidine | 5.307 | 6.33E-06 | 7.83E-05 |
| Histidine. N-tau-methyl- | 1.6573 | 0.1064 | 0.19565 |
| Hydroquinone | 1.553 | 0.12943 | 0.22947 |
| Hypotaurine | -1.6107 | 0.11624 | 0.21027 |
| Hypoxanthine | -1.5469 | 0.13088 | 0.2302 |
| Idose | -4.9733 | 1.74E-05 | 0.00016462 |
| Indole-3-acetamide | 2.8668 | 0.0069753 | 0.023164 |
| Indole-3-acetic acid | -2.186 | 0.035595 | 0.082071 |
| Indole. 3-hydroxy- | -1.2067 | 0.23565 | 0.35245 |
| Isobutanoic acid. 3-amino- | -3.2167 | 0.0027911 | 0.011008 |
| Isocaproic acid. 2-oxo | -0.83946 | 0.40691 | 0.53572 |
| Isoleucine | -3.9309 | 0.00038106 | 0.002261 |
| Isomaltose | -3.9777 | 0.00033298 | 0.0020298 |
| isopropyl beta-D-1-thiogalactopyranoside [19.097] | -2.0373 | 0.049234 | 0.10533 |
| Isovaleric acid. 2-oxo | -2.2275 | 0.032443 | 0.077205 |
| Lactic acid | 1.1947 | 0.24023 | 0.35753 |
| Lactic acid. 3-(4-hydroxyphenyl)- | -0.67357 | 0.50501 | 0.63672 |
| Loganin | -2.2906 | 0.028126 | 0.068769 |
| Lysine | -1.4131 | 0.16646 | 0.27847 |
| Malic acid | -4.4971 | 7.25E-05 | 0.00059765 |
| Malonic acid | -1.9016 | 0.065475 | 0.13365 |
| Mannopyranoside | -0.1157 | 0.90855 | 0.94685 |
| Methionine | -4.3378 | 0.00011627 | 0.00090771 |
| Methoxytryptamine | -1.8304 | 0.075722 | 0.14976 |
| Muramic acid. N-acetyl | 2.9688 | 0.0053658 | 0.019102 |
| Myo-Inositol | 2.1817 | 0.035939 | 0.082437 |
| N-acetyl Galactosamine | -3.7406 | 0.00065647 | 0.0034777 |
| N-acetyl- Aspartic acid | -6.0036 | 7.64E-07 | 1.51E-05 |
| N-Carboxyglycine | 4.3767 | 0.00010365 | 0.00082364 |
| N-methyl trans-4-hydroxy-L-proline (2S.4R)-4-hydroxy-1-methyl pyrrolidine-2-carboxylic acid | 2.3286 | 0.025782 | 0.064332 |
| Neryl acetate | -0.40074 | 0.69104 | 0.79267 |
| Neuraminic acid. N-glycolyl | -0.12492 | 0.9013 | 0.94685 |
| Nivalenol | 3.5493 | 0.0011237 | 0.0051553 |
| Nonanoic acid | 1.9751 | 0.056186 | 0.1185 |
| Norleucine | 1.1695 | 0.25011 | 0.36976 |
| Norvaline | 1.1294 | 0.26639 | 0.38614 |
| O-Toluic-acid | 0.984 | 0.33187 | 0.46295 |
| Octadecadienoic acid | 4.0546 | 0.00026649 | 0.0017264 |
| Octadecenoic acid | 2.0628 | 0.04661 | 0.10069 |
| Octadecenoic acid methyl ester | 6.2008 | 4.20E-07 | 1.17E-05 |
| Octanoic acid | 1.4712 | 0.15017 | 0.25701 |
| Ornithine | -3.3475 | 0.00196 | 0.0082088 |
| Oxalic acid | 0.1164 | 0.908 | 0.94685 |
| Pantothenic acid | 1.0473 | 0.30212 | 0.42816 |
| Pentanoic acid. 2-propy | -0.5757 | 0.5685 | 0.69693 |
| Phenol | 0.13564 | 0.89288 | 0.94154 |
| Phenylalanine | -6.7162 | 8.91E-08 | 3.30E-06 |
| Phosphoric acid | 0.9844 | 0.33168 | 0.46295 |
| Phosphoric acid monomethyl ester | 3.5914 | 0.00099922 | 0.0047812 |
| Proline | -1.3234 | 0.19429 | 0.30602 |
| Proline. 4-hydroxy | 1.7718 | 0.08513 | 0.16762 |
| Propane-1.2-diol | -1.9186 | 0.063226 | 0.13026 |
| Psicose | -1.0559 | 0.29824 | 0.42537 |
| Pyridine. 2-hydroxy- | 1.6771 | 0.10244 | 0.19073 |
| Pyridine. 3-hydroxy- | 2.1276 | 0.04049 | 0.089642 |
| Pyroglutamic acid | 0.14262 | 0.88741 | 0.938 |
| Pyruvic acid | -3.0805 | 0.0040081 | 0.01462 |
| Ribitol | -0.61739 | 0.54098 | 0.67057 |
| Ribonic acid | -2.3212 | 0.026225 | 0.064834 |
| Ribose | 0.020181 | 0.98401 | 0.98401 |
| Salicin | -2.7048 | 0.010485 | 0.031957 |
| Salicylic acid | -2.1731 | 0.036628 | 0.083161 |
| Sarcosine | 0.19105 | 0.84959 | 0.90481 |
| Sarcosineamide | 1.2164 | 0.23196 | 0.35168 |
| Sedoheptulose. 2.7-anhydro-. beta- | 1.8945 | 0.06644 | 0.135 |
| Serine | -0.94264 | 0.35233 | 0.48541 |
| Serine. O-phospho- | -0.080284 | 0.93647 | 0.9602 |
| Siloxane | 1.1419 | 0.26126 | 0.38097 |
| similar to Asparagine Derivate | 4.3033 | 0.00012872 | 0.00098432 |
| similar to Glycerolaldopyranosid | -2.3764 | 0.023087 | 0.060169 |
| similar to Pentasiloxane. dodecamethyl | -6.3921 | 2.36E-07 | 7.50E-06 |
| Sorbose | -0.71775 | 0.47767 | 0.60907 |
| Tartaric acid | 1.4625 | 0.15252 | 0.25906 |
| Tartronic acid | -1.5622 | 0.12725 | 0.22741 |
| Tetradecanoic acid | -1.1556 | 0.25568 | 0.3755 |
| Threonic acid-1.4-lactone | 2.1461 | 0.038874 | 0.087369 |
| Threonine | 1.2077 | 0.23526 | 0.35245 |
| Tryptophan | 2.6927 | 0.010801 | 0.032697 |
| Tyrosine | -2.499 | 0.017296 | 0.048408 |
| Tyrosine.1 | 0.35221 | 0.7268 | 0.81879 |
| Unknown01 | -1.5507 | 0.12996 | 0.2295 |
| Unknown02 | 5.0171 | 1.52E-05 | 0.00015068 |
| Unknown03 | 2.0181 | 0.051293 | 0.10921 |
| Unknown04 | 2.2206 | 0.032947 | 0.077987 |
| Unknown05 | 5.2795 | 6.88E-06 | 8.28E-05 |
| Unknown06 | 2.5662 | 0.014719 | 0.041719 |
| Unknown07 | 2.387 | 0.022522 | 0.060014 |
| Unknown08 | -5.4883 | 3.65E-06 | 4.95E-05 |
| Unknown09 | 1.5565 | 0.1286 | 0.2289 |
| Unknown10 | -2.4957 | 0.017432 | 0.048439 |
| Unknown100 | 2.3885 | 0.022443 | 0.060014 |
| Unknown101 | -0.054361 | 0.95696 | 0.96879 |
| Unknown102 | 1.3823 | 0.17565 | 0.29036 |
| Unknown103 | -0.05706 | 0.95482 | 0.96879 |
| Unknown104 | -4.0477 | 0.00027189 | 0.0017264 |
| Unknown105 | -4.9251 | 2.01E-05 | 0.00018263 |
| Unknown106 | 4.2429 | 0.00015378 | 0.0011405 |
| Unknown107 | 1.062 | 0.29553 | 0.42287 |
| Unknown108 | 9.87 | 1.19E-11 | 2.65E-09 |
| Unknown109 | 2.2749 | 0.029146 | 0.070565 |
| Unknown11 | -0.26395 | 0.79337 | 0.87388 |
| Unknown110 | -5.1154 | 1.13E-05 | 0.00012283 |
| Unknown111 | -5.2414 | 7.73E-06 | 9.05E-05 |
| Unknown112 | -5.2144 | 8.39E-06 | 9.33E-05 |
| Unknown113 | 1.4599 | 0.15325 | 0.2593 |
| Unknown114 | 0.3789 | 0.70705 | 0.80264 |
| Unknown115 | -4.9375 | 1.94E-05 | 0.0001796 |
| Unknown116 | -3.3449 | 0.0019738 | 0.0082088 |
| Unknown117 | -3.7295 | 0.00067738 | 0.0035463 |
| Unknown118 | 1.1672 | 0.25102 | 0.36987 |
| Unknown119 | 1.3805 | 0.17617 | 0.29036 |
| Unknown12 | 5.8586 | 1.19E-06 | 2.03E-05 |
| Unknown120 | 1.9324 | 0.061439 | 0.12776 |
| Unknown121 | 0.39157 | 0.69775 | 0.79414 |
| Unknown122 | -0.25517 | 0.80008 | 0.87694 |
| Unknown123 | -6.1171 | 5.41E-07 | 1.27E-05 |
| Unknown124 | -5.4281 | 4.38E-06 | 5.74E-05 |
| Unknown125 | 1.6689 | 0.10406 | 0.19214 |
| Unknown126 | -1.4944 | 0.14404 | 0.25038 |
| Unknown127 | -2.2531 | 0.030626 | 0.073271 |
| Unknown128 | 0.23855 | 0.81285 | 0.88224 |
| Unknown129 | -0.93924 | 0.35405 | 0.48559 |
| Unknown13 | 5.7425 | 1.69E-06 | 2.59E-05 |
| Unknown130 | -0.29944 | 0.76637 | 0.85046 |
| Unknown131 | -0.8908 | 0.37912 | 0.50816 |
| Unknown132 | 1.4992 | 0.1428 | 0.2492 |
| Unknown133 | 2.3752 | 0.023149 | 0.060169 |
| Unknown134 | 1.8595 | 0.071376 | 0.14307 |
| Unknown135 | -6.1164 | 5.43E-07 | 1.27E-05 |
| Unknown136 | 4.0086 | 0.00030454 | 0.0018822 |
| Unknown137 | 2.9642 | 0.0054304 | 0.019179 |
| Unknown138 | -0.62665 | 0.53495 | 0.66496 |
| Unknown139 | 0.32502 | 0.7471 | 0.83743 |
| Unknown14 | 2.3499 | 0.024544 | 0.062771 |
| Unknown140 | 3.9093 | 0.00040551 | 0.0023435 |
| Unknown141 | 1.3192 | 0.19567 | 0.30602 |
| Unknown142 | 0.53988 | 0.5927 | 0.71284 |
| Unknown143 | -1.05 | 0.30092 | 0.42783 |
| Unknown144 | -1.216 | 0.2321 | 0.35168 |
| Unknown145 | -0.1958 | 0.8459 | 0.90481 |
| Unknown146 | -2.1868 | 0.035529 | 0.082071 |
| Unknown147 | 0.95227 | 0.34749 | 0.48172 |
| Unknown148 | -0.84114 | 0.40598 | 0.53572 |
| Unknown149 | 2.0852 | 0.044409 | 0.0964 |
| Unknown15 | 3.6478 | 0.00085315 | 0.0043142 |
| Unknown150 | -0.50342 | 0.61783 | 0.73315 |
| Unknown151 | -1.5743 | 0.1244 | 0.22323 |
| Unknown152 | -0.30235 | 0.76418 | 0.85015 |
| Unknown153 | -2.0114 | 0.052033 | 0.11026 |
| Unknown154 | -0.88661 | 0.38134 | 0.5096 |
| Unknown155 | -1.854 | 0.072186 | 0.14405 |
| Unknown156 | 0.31538 | 0.75435 | 0.84343 |
| Unknown157 | -1.9506 | 0.059143 | 0.12356 |
| Unknown158 | -0.91038 | 0.36885 | 0.50195 |
| Unknown159 | -1.5355 | 0.13364 | 0.23414 |
| Unknown16 | -1.3214 | 0.19495 | 0.30602 |
| Unknown160 | -2.8472 | 0.0073319 | 0.024168 |
| Unknown161 | 0.16451 | 0.87027 | 0.92428 |
| Unknown162 | -5.0573 | 1.35E-05 | 0.00013963 |
| Unknown163 | -1.3241 | 0.19406 | 0.30602 |
| Unknown164 | 0.67345 | 0.50509 | 0.63672 |
| Unknown165 | -2.383 | 0.022732 | 0.060032 |
| Unknown166 | -1.3209 | 0.19509 | 0.30602 |
| Unknown167 | -4.5735 | 5.78E-05 | 0.00049434 |
| Unknown168 | -3.552 | 0.0011154 | 0.0051553 |
| Unknown169 | 0.024838 | 0.98032 | 0.98401 |
| Unknown17 | 1.6932 | 0.099292 | 0.18666 |
| Unknown170 | -29.504 | 2.57E-26 | 1.15E-23 |
| Unknown171 | -1.1533 | 0.25661 | 0.37563 |
| Unknown172 | 1.3754 | 0.17774 | 0.29076 |
| Unknown173 | 2.5672 | 0.014685 | 0.041719 |
| Unknown174 | 2.3336 | 0.025488 | 0.06408 |
| Unknown175 | 2.2084 | 0.033859 | 0.079722 |
| Unknown176 | -0.8996 | 0.37448 | 0.50549 |
| Unknown177 | 2.2718 | 0.029356 | 0.070614 |
| Unknown178 | 2.2745 | 0.029177 | 0.070565 |
| Unknown179 | -0.67713 | 0.50278 | 0.63672 |
| Unknown18 | -6.021 | 7.24E-07 | 1.51E-05 |
| Unknown180 | -1.6926 | 0.099413 | 0.18666 |
| Unknown181 | 1.7544 | 0.088111 | 0.17273 |
| Unknown182 | 1.484 | 0.14677 | 0.25315 |
| Unknown183 | 1.2435 | 0.22195 | 0.34038 |
| Unknown184 | 0.81647 | 0.41975 | 0.54938 |
| Unknown185 | 2.7311 | 0.0098188 | 0.030555 |
| Unknown186 | 2.739 | 0.0096275 | 0.030385 |
| Unknown187 | -7.5814 | 6.90E-09 | 5.12E-07 |
| Unknown188 | -3.1036 | 0.0037717 | 0.014182 |
| Unknown189 | -2.3516 | 0.024451 | 0.062771 |
| Unknown19 | 3.637 | 0.00087945 | 0.0043484 |
| Unknown190 | -4.5259 | 6.66E-05 | 0.00055884 |
| Unknown191 | 1.6272 | 0.11267 | 0.20549 |
| Unknown192 | -3.7741 | 0.00059695 | 0.0033205 |
| Unknown193 | 2.7527 | 0.0093055 | 0.029578 |
| Unknown194 | 1.5985 | 0.11893 | 0.21427 |
| Unknown195 | -6.2281 | 3.87E-07 | 1.15E-05 |
| Unknown196 | 0.54209 | 0.59119 | 0.71284 |
| Unknown197 | -0.51128 | 0.61236 | 0.72861 |
| Unknown198 | 0.053159 | 0.95791 | 0.96879 |
| Unknown199 | -0.35853 | 0.7221 | 0.81557 |
| Unknown20 | -0.11789 | 0.90683 | 0.94685 |
| Unknown200 | -7.0249 | 3.55E-08 | 1.58E-06 |
| Unknown201 | 0.11872 | 0.90618 | 0.94685 |
| Unknown202 | 3.6282 | 0.0009014 | 0.00436 |
| Unknown203 | -0.094267 | 0.92544 | 0.95328 |
| Unknown204 | -2.3922 | 0.022253 | 0.060014 |
| Unknown205 | -1.3215 | 0.19492 | 0.30602 |
| Unknown206 | -1.468 | 0.15104 | 0.25751 |
| Unknown207 | 0.22232 | 0.82536 | 0.89364 |
| Unknown208 | 0.52445 | 0.60327 | 0.7236 |
| Unknown209 | 0.070414 | 0.94427 | 0.96376 |
| Unknown21 | 2.6729 | 0.011344 | 0.033654 |
| Unknown210 | 2.3436 | 0.024906 | 0.062973 |
| Unknown211 | -1.6298 | 0.1121 | 0.20529 |
| Unknown212 | 8.0286 | 1.89E-09 | 2.11E-07 |
| Unknown213 | 3.7673 | 0.00060851 | 0.003343 |
| Unknown214 | -0.64238 | 0.52481 | 0.65601 |
| Unknown215 | 4.0432 | 0.00027545 | 0.0017264 |
| Unknown216 | 1.3827 | 0.17552 | 0.29036 |
| Unknown217 | -1.3712 | 0.17903 | 0.29076 |
| Unknown218 | -0.78399 | 0.43832 | 0.56701 |
| Unknown219 | 0.23968 | 0.81197 | 0.88224 |
| Unknown22 | -2.0896 | 0.043992 | 0.095964 |
| Unknown220 | 4.1951 | 0.00017691 | 0.0012496 |
| Unknown221 | -3.5532 | 0.0011117 | 0.0051553 |
| Unknown222 | -0.1115 | 0.91186 | 0.94808 |
| Unknown223 | 2.1861 | 0.035589 | 0.082071 |
| Unknown224 | 0.75452 | 0.45559 | 0.58258 |
| Unknown225 | 2.6369 | 0.012394 | 0.036526 |
| Unknown226 | 0.57008 | 0.57226 | 0.69769 |
| Unknown227 | 0.63196 | 0.53152 | 0.66254 |
| Unknown228 | 3.3372 | 0.0020154 | 0.008304 |
| Unknown229 | -0.10419 | 0.91761 | 0.94962 |
| Unknown23 | 0.8574 | 0.39706 | 0.52901 |
| Unknown230 | 2.9051 | 0.0063235 | 0.021646 |
| Unknown231 | 0.32899 | 0.74413 | 0.8362 |
| Unknown232 | 0.97667 | 0.33543 | 0.46646 |
| Unknown233 | -0.57034 | 0.57209 | 0.69769 |
| Unknown234 | 0.84441 | 0.40417 | 0.53572 |
| Unknown235 | 0.45104 | 0.65474 | 0.76073 |
| Unknown236 | -0.5535 | 0.58344 | 0.70552 |
| Unknown237 | 1.2898 | 0.20559 | 0.31988 |
| Unknown238 | 2.0603 | 0.046865 | 0.10075 |
| Unknown239 | 0.67765 | 0.50245 | 0.63672 |
| Unknown24 | -1.747 | 0.089417 | 0.17366 |
| Unknown240 | -0.2055 | 0.83837 | 0.90115 |
| Unknown241 | 0.81358 | 0.42138 | 0.5499 |
| Unknown242 | 0.23893 | 0.81256 | 0.88224 |
| Unknown243 | -0.10608 | 0.91613 | 0.94962 |
| Unknown244 | -1.3385 | 0.18938 | 0.30314 |
| Unknown245 | -0.15379 | 0.87866 | 0.93096 |
| Unknown246 | -2.3732 | 0.023256 | 0.060169 |
| Unknown247 | -2.7123 | 0.010291 | 0.031581 |
| Unknown248 | -5.5681 | 2.87E-06 | 4.11E-05 |
| Unknown249 | 3.0175 | 0.0047278 | 0.016967 |
| Unknown25 | 1.122 | 0.2695 | 0.38938 |
| Unknown250 | 0.75803 | 0.45351 | 0.58238 |
| Unknown251 | -2.8414 | 0.0074403 | 0.024345 |
| Unknown252 | -3.3964 | 0.0017148 | 0.0073375 |
| Unknown253 | 3.6432 | 0.00086432 | 0.0043216 |
| Unknown254 | 5.0404 | 1.42E-05 | 0.00014361 |
| Unknown255 | 2.6743 | 0.011306 | 0.033654 |
| Unknown256 | 0.26823 | 0.79009 | 0.87244 |
| Unknown257 | 2.3818 | 0.022799 | 0.060032 |
| Unknown258 | 0.89883 | 0.37489 | 0.50549 |
| Unknown259 | 4.7129 | 3.81E-05 | 0.00033215 |
| Unknown26 | -1.708 | 0.096488 | 0.18428 |
| Unknown260 | 1.36 | 0.18252 | 0.29427 |
| Unknown261 | 4.858 | 2.46E-05 | 0.00021913 |
| Unknown262 | -0.49218 | 0.62567 | 0.74048 |
| Unknown263 | 2.589 | 0.01393 | 0.040138 |
| Unknown264 | -0.45476 | 0.65209 | 0.76073 |
| Unknown265 | -0.66409 | 0.51099 | 0.6414 |
| Unknown266 | 2.4457 | 0.019627 | 0.053583 |
| Unknown267 | 1.2154 | 0.23234 | 0.35168 |
| Unknown268 | 2.1167 | 0.041469 | 0.090905 |
| Unknown269 | -1.6879 | 0.10032 | 0.18757 |
| Unknown27 | 1.4024 | 0.1696 | 0.28267 |
| Unknown270 | -1.6959 | 0.098779 | 0.18666 |
| Unknown271 | -2.128 | 0.040452 | 0.089642 |
| Unknown272 | -0.40062 | 0.69113 | 0.79267 |
| Unknown273 | 3.0921 | 0.0038881 | 0.014418 |
| Unknown274 | -2.1755 | 0.036429 | 0.083133 |
| Unknown275 | 0.43673 | 0.66499 | 0.77062 |
| Unknown276 | -0.20926 | 0.83546 | 0.90019 |
| Unknown277 | -0.94695 | 0.35016 | 0.48391 |
| Unknown278 | -3.1947 | 0.0029607 | 0.011557 |
| Unknown279 | 3.5247 | 0.0012035 | 0.0054098 |
| Unknown28 | -3.0867 | 0.0039437 | 0.014504 |
| Unknown280 | -3.2162 | 0.0027952 | 0.011008 |
| Unknown281 | -3.7778 | 0.00059064 | 0.0033205 |
| Unknown282 | -1.1714 | 0.24935 | 0.36976 |
| Unknown283 | -5.8324 | 1.28E-06 | 2.12E-05 |
| Unknown284 | -2.6816 | 0.011103 | 0.033384 |
| Unknown285 | -4.2332 | 0.00015817 | 0.0011539 |
| Unknown286 | -3.4036 | 0.0016812 | 0.0073344 |
| Unknown288 | -0.9943 | 0.3269 | 0.4589 |
| Unknown289 | 0.51903 | 0.607 | 0.72612 |
| Unknown29 | -1.2057 | 0.23602 | 0.35245 |
| Unknown290 | 5.9596 | 8.73E-07 | 1.62E-05 |
| Unknown291 | -5.6034 | 2.57E-06 | 3.82E-05 |
| Unknown292 | 6.9162 | 4.90E-08 | 1.98E-06 |
| Unknown293 | -2.8327 | 0.0076058 | 0.024705 |
| Unknown294 | 0.085101 | 0.93267 | 0.95851 |
| Unknown295 | -1.8922 | 0.06676 | 0.13504 |
| Unknown296 | -1.2571 | 0.21704 | 0.33536 |
| Unknown30 | -0.45496 | 0.65194 | 0.76073 |
| Unknown31 | -0.78926 | 0.43527 | 0.56636 |
| Unknown32 | 5.9318 | 9.50E-07 | 1.69E-05 |
| Unknown33 | -0.90502 | 0.37164 | 0.50421 |
| Unknown34 | 2.4935 | 0.017525 | 0.048439 |
| Unknown35 | 3.6674 | 0.00080735 | 0.0041295 |
| Unknown36 | 0.663 | 0.51168 | 0.6414 |
| Unknown37 | -4.2986 | 0.0001305 | 0.00098432 |
| Unknown38 | 2.8769 | 0.0067969 | 0.022914 |
| Unknown39 | -2.154 | 0.038206 | 0.086302 |
| Unknown40 | -2.9092 | 0.0062585 | 0.021589 |
| Unknown41 | -4.2082 | 0.00017022 | 0.0012218 |
| Unknown42 | -3.1407 | 0.003419 | 0.013004 |
| Unknown43 | -0.56538 | 0.57543 | 0.69963 |
| Unknown44 | -1.7016 | 0.097695 | 0.18579 |
| Unknown45 | 7.3878 | 1.22E-08 | 7.73E-07 |
| Unknown46 | 7.2025 | 2.10E-08 | 1.17E-06 |
| Unknown47 | -0.25537 | 0.79993 | 0.87694 |
| Unknown48 | -0.91136 | 0.36834 | 0.50195 |
| Unknown49 | -0.77959 | 0.44087 | 0.56866 |
| Unknown50 | -1.6189 | 0.11444 | 0.20786 |
| Unknown51 | 0.30778 | 0.76007 | 0.8477 |
| Unknown52 | 0.51448 | 0.61015 | 0.72793 |
| Unknown53 | -2.484 | 0.017928 | 0.049246 |
| Unknown54 | 1.3182 | 0.19599 | 0.30602 |
| Unknown55 | 2.8731 | 0.0068633 | 0.022964 |
| Unknown56 | 6.1476 | 4.94E-07 | 1.27E-05 |
| Unknown57 | 0.48056 | 0.63382 | 0.74814 |
| Unknown58 | -2.6072 | 0.013328 | 0.038763 |
| Unknown59 | -3.4336 | 0.0015484 | 0.0068549 |
| Unknown60 | -1.9052 | 0.065001 | 0.1333 |
| Unknown61 | -1.1401 | 0.26197 | 0.38097 |
| Unknown62 | -0.37295 | 0.71143 | 0.80557 |
| Unknown63 | 0.40729 | 0.68628 | 0.79118 |
| Unknown64 | -3.5253 | 0.0012015 | 0.0054098 |
| Unknown65 | 1.6729 | 0.10326 | 0.19146 |
| Unknown66 | -0.25185 | 0.80263 | 0.87757 |
| Unknown67 | -3.7077 | 0.00072053 | 0.0037283 |
| Unknown68 | 0.044447 | 0.9648 | 0.97355 |
| Unknown69 | -2.762 | 0.0090905 | 0.029103 |
| Unknown70 | -1.3671 | 0.1803 | 0.29176 |
| Unknown71 | 5.4868 | 3.67E-06 | 4.95E-05 |
| Unknown72 | 2.5552 | 0.015115 | 0.04257 |
| Unknown73 | -0.60758 | 0.54739 | 0.67476 |
| Unknown74 | 4.1342 | 0.0002114 | 0.0014253 |
| Unknown75 | 0.81963 | 0.41797 | 0.54867 |
| Unknown76 | 0.89672 | 0.376 | 0.50549 |
| Unknown77 | 2.327 | 0.025877 | 0.064332 |
| Unknown78 | 0.071523 | 0.94339 | 0.96376 |
| Unknown79 | -3.0555 | 0.0042812 | 0.015489 |
| Unknown80 | 0.39367 | 0.69621 | 0.79414 |
| Unknown81 | -3.2896 | 0.0022932 | 0.0091935 |
| Unknown82 | -0.45167 | 0.65429 | 0.76073 |
| Unknown83 | -2.7158 | 0.010202 | 0.031526 |
| Unknown84 | 3.954 | 0.00035653 | 0.002144 |
| Unknown85 | 1.0068 | 0.32095 | 0.45196 |
| Unknown86 | 0.75699 | 0.45412 | 0.58238 |
| Unknown87 | 0.46718 | 0.64327 | 0.75728 |
| Unknown88 | 0.025196 | 0.98004 | 0.98401 |
| Unknown89 | 5.4042 | 4.71E-06 | 5.99E-05 |
| Unknown90 | -0.021737 | 0.98278 | 0.98401 |
| Unknown91 | 4.0522 | 0.00026834 | 0.0017264 |
| Unknown92 | -3.7554 | 0.00062946 | 0.0034048 |
| Unknown93 | -6.527 | 1.57E-07 | 5.38E-06 |
| Unknown94 | 7.6322 | 5.95E-09 | 5.12E-07 |
| Unknown95 | -5.9959 | 7.82E-07 | 1.51E-05 |
| Unknown96 | -5.7774 | 1.52E-06 | 2.41E-05 |
| Unknown97 | 0.93805 | 0.35465 | 0.48559 |
| Unknown98 | 3.1608 | 0.0032409 | 0.012433 |
| Unknown99 | 2.9376 | 0.0058177 | 0.020225 |
| Urea | -2.891 | 0.0065564 | 0.022272 |
| Uric acid | 0.56116 | 0.57827 | 0.70117 |
| Valeric acid. 2-oxo- | -1.8791 | 0.068584 | 0.1381 |
| Valine | -3.1706 | 0.0031572 | 0.012217 |
| Xanthine | -1.0189 | 0.31524 | 0.44534 |
| Xylitol | 0.21841 | 0.82838 | 0.89473 |

**Table S3.** Pairwise comparison of metabolites between HF-Frail and HF without physical frailty.

|  | t.stat | p.value |
| --- | --- | --- |
| 2-aminobutanoic acid | 0.99611 | 0.33003 |
| 2-Furoic acid | -1.4479 | 0.16175 |
| 2-hydroxybutanoic acid | -0.39019 | 0.70015 |
| 2-Piperidinecarboxamide | 1.8522 | 0.077464 |
| 2-Piperidinecarboxamide.1 | 0.16374 | 0.87143 |
| 2-PIPERIDINECARBOXYLIC ACID | 1.1341 | 0.26895 |
| 2,4-Dihydroxybutanoic acid | -1.5287 | 0.14059 |
| 3-Hydroxybutyric acid | 0.73574 | 0.46966 |
| 3-Methyl-2-oxovaleric acid | -0.007728 | 0.9939 |
| 4-hydroxyphenylacetic acid | -0.83871 | 0.41065 |
| Adipic acid. 2-amino | -0.51611 | 0.61093 |
| Alanine | 0.99656 | 0.32982 |
| alanine | 0.80597 | 0.42889 |
| Androstenedione | 1.5945 | 0.12509 |
| Arabinoheptulosonic acid enol. 3-deoxy | -0.41149 | 0.68469 |
| Arabitol | -0.41703 | 0.6807 |
| Asparagine | 0.040459 | 0.96809 |
| Asparagine Derivate | -0.25254 | 0.80297 |
| Aspartic acid | -1.3914 | 0.17801 |
| Benzoic acid | 0.43704 | 0.66635 |
| Benzylalcohol | 0.037945 | 0.97007 |
| beta-D-allose | -0.065828 | 0.94811 |
| Caffeine | 0.3881 | 0.70167 |
| Capric Acid | -0.74364 | 0.46497 |
| Carbodiimides | 0.29984 | 0.76711 |
| Catechol | -0.11679 | 0.90809 |
| Citric acid | 0.57705 | 0.56976 |
| Creatinine | -0.92904 | 0.36295 |
| Cystamine | 1.7483 | 0.094359 |
| Cysteine | -0.088923 | 0.92995 |
| Cysteinylglycine | -1.2369 | 0.22915 |
| Cystine | 1.4154 | 0.17096 |
| D-Erythrose | 1.3017 | 0.20649 |
| Diethylene glycol | 0.56872 | 0.57531 |
| dihydrotestosterone1 | 1.6607 | 0.11096 |
| dihydrotestosterone2 | 0.28053 | 0.78169 |
| Ergothioneine | 0.57494 | 0.57117 |
| Erythritol | 1.4686 | 0.15611 |
| Fructose | -1.2135 | 0.2378 |
| Fructose.1 | -1.8759 | 0.073997 |
| Fucose | 1.0781 | 0.29265 |
| Fumaric acid | -0.96761 | 0.34376 |
| Galactonic acid | -1.0963 | 0.28479 |
| Galactonic acid 1,4-lactone, tetra | 0.37377 | 0.71215 |
| Galactose | -0.57536 | 0.57089 |
| Galactose. 2-amino-2-deoxy | -0.5379 | 0.59605 |
| Galactose. 2-deoxy | -1.9706 | 0.061479 |
| Galacturonic acid | -1.1967 | 0.24415 |
| Galacturonic acid-1-phosphate | 2.2455 | 0.035112 |
| GalNAc-ol | -0.21093 | 0.83488 |
| Glucoheptonic Acid | 1.1571 | 0.25965 |
| Gluconic acid-1.4-lactone | -0.1075 | 0.91536 |
| Glucopyranose | 0.035276 | 0.97218 |
| Glucose. 2-deoxy | -0.10617 | 0.91641 |
| Glutamic acid | 2.1517 | 0.042662 |
| Glutamic acid.1 | -2.4197 | 0.024247 |
| Glutamine | -0.80483 | 0.42953 |
| Glutaric acid | -0.24897 | 0.80569 |
| Glutaric acid. 2-hydroxy | -1.9239 | 0.067402 |
| Glutaric acid. 2-oxo | -1.0249 | 0.31655 |
| Glyceric acid | -0.82577 | 0.4178 |
| Glycerol | -0.40809 | 0.68715 |
| Glycerolaldopyranosid | -1.1968 | 0.24414 |
| Glycine | 0.6596 | 0.51636 |
| Glycolic acid | -0.10546 | 0.91697 |
| Glyoxylic acid | 0.82138 | 0.42024 |
| Heptadecanoic acid | 0.93641 | 0.35923 |
| Hexadecenoic acid | -0.27792 | 0.78367 |
| Hippuric acid | 0.099853 | 0.92137 |
| Histidine | 0.82678 | 0.41724 |
| Histidine. N-tau-methyl | 1.0338 | 0.31245 |
| Hydroquinone | 0.58768 | 0.56273 |
| Hypotaurine | 0.17589 | 0.86199 |
| Hypoxanthine | 0.26989 | 0.78976 |
| Idose | -1.2439 | 0.22664 |
| Indole-3-acetamide | 2.4189 | 0.024286 |
| Indole-3-acetic acid | -1.7817 | 0.088604 |
| Indole. 3-hydroxy | 0.40212 | 0.69148 |
| Isobutanoic acid. 3-amino | 0.32395 | 0.74904 |
| Isocaproic acid. 2-oxo | 1.4156 | 0.17091 |
| Isoleucine | 2.0093 | 0.056934 |
| ISOMALTOSE | 0.97221 | 0.34151 |
| Isopropyl beta-D-1-thiogalactopyranoside | -0.57187 | 0.57321 |
| Isovaleric acid. 2-oxo | -0.054438 | 0.95708 |
| Lactic acid | 0.38206 | 0.70608 |
| Lactic acid. 3-(4-hydroxyphenyl) | 1.0374 | 0.31083 |
| Lauric Acid | -0.96535 | 0.34486 |
| Loganin | -0.66621 | 0.5122 |
| Lysine | 0.056955 | 0.9551 |
| Malic acid | -1.4282 | 0.16727 |
| Malonic acid | 0.46054 | 0.64965 |
| Mannopyranoside. 1-O-methyl-. Alpha | -1.3326 | 0.19629 |
| Methionine | 2.1372 | 0.04395 |
| Methoxytryptamine | -0.099062 | 0.92199 |
| Methylcysteine | 1.2425 | 0.22712 |
| Muramic acid | 0.066766 | 0.94737 |
| Myo-Inositol | -0.070684 | 0.94429 |
| N-Acetyl-L-Aspartic Acid | 1.5696 | 0.13078 |
| N-Carboxyglycine | -0.59358 | 0.55885 |
| N-methyl trans-4-hydroxy-L-proline (2S.4R)-4-hydroxy-1-methyl pyrrolidine-2-carboxylic acid | 0.16825 | 0.86792 |
| Neryl acetate | -0.29397 | 0.77154 |
| Neuraminic acid. N-glycolyl | 0.93197 | 0.36147 |
| Nivalenol | -2.5684 | 0.01753 |
| Nonanoic acid | 1.4203 | 0.16954 |
| Norleucine | 0.012032 | 0.99051 |
| Norvaline. | 0.27491 | 0.78595 |
| O-Toluic-acid | -0.44504 | 0.66064 |
| Octadecadienoic acid | -0.14112 | 0.88906 |
| Octadecenoic acid | -1.0656 | 0.29818 |
| Octadecenoic acid methyl ester | 1.666 | 0.10989 |
| Octanoic acid | 0.25479 | 0.80125 |
| Ornithine | 1.2768 | 0.21497 |
| Oxalic acid | 0.18167 | 0.8575 |
| Pantothenic acid (Vitamin B5) | 0.92877 | 0.36309 |
| Pentanoic acid. 2-propyl | -0.57126 | 0.57361 |
| Pentasiloxane. dodecamethyl | 0.60514 | 0.55128 |
| Phenol | 0.13196 | 0.89622 |
| Phenylalanine | 0.029728 | 0.97655 |
| Phosphoric acid | -0.18571 | 0.85437 |
| Phosphoric acid monomethyl ester | 0.42724 | 0.67335 |
| Proline | -1.3213 | 0.19998 |
| Proline. 4-hydroxy | -1.4588 | 0.15876 |
| Propane-1.2-diol | -1.3549 | 0.18921 |
| Psicose | -0.10467 | 0.91759 |
| Pyridine. 2-hydroxy | 0.58856 | 0.56215 |
| Pyridine. 3-hydroxy | -0.12134 | 0.90452 |
| Pyroglutamic acid | 1.8787 | 0.073604 |
| Pyruvic acid | -1.6338 | 0.11653 |
| Ribitol | -1.4681 | 0.15624 |
| Ribonic acid | -0.36333 | 0.71983 |
| Ribose | -1.8532 | 0.077315 |
| Salicin | -1.1043 | 0.28138 |
| Salicylic acid | -1.602 | 0.12342 |
| Sarcosine | -1.0151 | 0.32111 |
| Sarcosineamide | 1.3246 | 0.1989 |
| Sedoheptulose. 2.7-anhydro- beta | -1.1072 | 0.28016 |
| Serine | -0.81295 | 0.42496 |
| Serine. O-phospho | -0.405 | 0.68939 |
| Siloxane | 0.44922 | 0.65766 |
| Sorbose | 0.25203 | 0.80336 |
| Tartaric acid | -0.39712 | 0.69511 |
| Tartronic acid | 0.3896 | 0.70058 |
| Tetradecanoic acid | -0.70966 | 0.48537 |
| Threonic acid-1.4-lactone | 0.85214 | 0.40332 |
| Threonine | -0.045723 | 0.96394 |
| Tryptophan | 1.7734 | 0.090013 |
| Tyrosine | 1.1093 | 0.27928 |
| Urea | 1.3975 | 0.17621 |
| Uric acid | 0.55115 | 0.58708 |
| Valeric acid. 2-oxo | 0.69342 | 0.4953 |
| Valine | 1.7642 | 0.091576 |
| Xanthine | 1.593 | 0.12542 |
| Xylitol | -1.3358 | 0.19526 |
| Unknown.1 | -1.3424 | 0.19317 |
| Unknown.10 | 0.048818 | 0.96151 |
| Unknown.11 | 0.10175 | 0.91987 |
| Unknown.12 | 0.050166 | 0.96044 |
| Unknown.13 | -0.84001 | 0.40994 |
| Unknown.14 | 0.48051 | 0.63561 |
| Unknown.2 | 1.386 | 0.17962 |
| Unknown.3 | 1.1791 | 0.25096 |
| Unknown.4 | 0.30408 | 0.76392 |
| Unknown.5 | 0.57577 | 0.57062 |
| Unknown.6 | -1.2702 | 0.21728 |
| Unknown.7 | -0.49066 | 0.62853 |
| Unknown.8 | 1.2521 | 0.22368 |
| Unknown.9 | -0.15068 | 0.8816 |
| Unknown#bth-pae-001 | 0.69126 | 0.49664 |
| Unknown#bth-pae-010 | -0.19255 | 0.84907 |
| Unknown#bth-pae-011 | 1.6932 | 0.10452 |
| Unknown#bth-pae-038 | 1.3415 | 0.19344 |
| Unknown#sst-cgl-010a | 0.75037 | 0.46098 |
| Unknown#sst-cgl-D05 | -2.3253 | 0.029672 |
| Unknown01 | 0.24059 | 0.8121 |
| Unknown02 | 0.56694 | 0.57649 |
| Unknown03 | -0.39206 | 0.69878 |
| Unknown04 | -0.053871 | 0.95752 |
| Unknown05 | -0.58743 | 0.5629 |
| Unknown06 | -1.391 | 0.17813 |
| Unknown07 | 0.50951 | 0.61547 |
| Unknown08 | -1.0261 | 0.31601 |
| Unknown09 | -0.098192 | 0.92267 |
| Unknown10 | 1.5518 | 0.13498 |
| Unknown100 | 0.17977 | 0.85898 |
| Unknown101 | 1.5114 | 0.14492 |
| Unknown102 | -0.44207 | 0.66275 |
| Unknown103 | 0.56359 | 0.57873 |
| Unknown104 | 0.21844 | 0.8291 |
| Unknown105 | -2.7186 | 0.012544 |
| Unknown106 | 1.3681 | 0.18508 |
| Unknown107 | -0.86624 | 0.39571 |
| Unknown108 | -1.6858 | 0.10597 |
| Unknown109 | 0.79611 | 0.43448 |
| Unknown11 | 0.077122 | 0.93922 |
| Unknown110 | -0.52588 | 0.60423 |
| Unknown111 | 0.72065 | 0.47871 |
| Unknown112 | 1.1968 | 0.24413 |
| Unknown113 | 0.63041 | 0.53492 |
| Unknown114 | 0.71413 | 0.48265 |
| Unknown115 | -0.28247 | 0.78022 |
| Unknown116 | 1.1828 | 0.24953 |
| Unknown117 | -0.83434 | 0.41306 |
| Unknown118 | -0.65563 | 0.51886 |
| Unknown119 | -2.4821 | 0.021177 |
| Unknown12 | 0.026926 | 0.97876 |
| Unknown120 | 0.33088 | 0.74387 |
| Unknown121 | -0.70581 | 0.48772 |
| Unknown122 | 0.017498 | 0.9862 |
| Unknown123 | 0.44917 | 0.6577 |
| Unknown124 | -1.392 | 0.17782 |
| Unknown125 | -0.50612 | 0.61781 |
| Unknown126 | 1.539 | 0.13807 |
| Unknown127 | 1.1259 | 0.27232 |
| Unknown128 | 0.92426 | 0.36538 |
| Unknown129 | -1.4751 | 0.15435 |
| Unknown13 | -0.42786 | 0.67291 |
| Unknown130 | 0.55923 | 0.58165 |
| Unknown131 | 0.15893 | 0.87518 |
| Unknown132 | -0.4551 | 0.65349 |
| Unknown133 | -0.91235 | 0.37147 |
| Unknown134 | -0.42781 | 0.67295 |
| Unknown135 | -0.45346 | 0.65466 |
| Unknown136 | -1.1931 | 0.24554 |
| Unknown137 | -1.103 | 0.28196 |
| Unknown138 | 0.21415 | 0.8324 |
| Unknown139 | -0.73305 | 0.47126 |
| Unknown14 | -0.04422 | 0.96513 |
| Unknown140 | 0.56361 | 0.57872 |
| Unknown141 | 2.3675 | 0.027124 |
| Unknown142 | 1.7792 | 0.089028 |
| Unknown143 | -0.86851 | 0.39449 |
| Unknown144 | -1.1536 | 0.26104 |
| Unknown145 | 1.3812 | 0.18107 |
| Unknown146 | 0.33437 | 0.74126 |
| Unknown147 | -1.9675 | 0.061859 |
| Unknown148 | 0.28825 | 0.77585 |
| Unknown149 | 0.77235 | 0.44813 |
| Unknown15 | -0.49421 | 0.62606 |
| Unknown150 | 1.1289 | 0.27109 |
| Unknown151 | 0.27124 | 0.78874 |
| Unknown152 | 0.080337 | 0.9367 |
| Unknown153 | 0.28014 | 0.78198 |
| Unknown154 | 0.5413 | 0.59374 |
| Unknown155 | -0.47928 | 0.63647 |
| Unknown156 | -1.1602 | 0.25838 |
| Unknown157 | -1.166 | 0.25611 |
| Unknown158 | -0.18342 | 0.85615 |
| Unknown159 | -0.53756 | 0.59628 |
| Unknown16 | 0.41446 | 0.68255 |
| Unknown160 | 1.4579 | 0.15899 |
| Unknown161 | 1.373 | 0.1836 |
| Unknown162 | -0.92701 | 0.36398 |
| Unknown163 | -0.86931 | 0.39406 |
| Unknown164 | -0.43017 | 0.67125 |
| Unknown165 | -0.7407 | 0.46671 |
| Unknown166 | -0.33022 | 0.74436 |
| Unknown167 | -1.1698 | 0.25459 |
| Unknown168 | -1.1576 | 0.25944 |
| Unknown169 | -0.41417 | 0.68276 |
| Unknown17 | 0.45403 | 0.65426 |
| Unknown170 | -0.64152 | 0.52781 |
| Unknown171 | 0.1284 | 0.899 |
| Unknown172 | 2.0324 | 0.054359 |
| Unknown173 | 0.026301 | 0.97925 |
| Unknown174 | 0.44405 | 0.66135 |
| Unknown175 | 0.32921 | 0.74511 |
| Unknown176 | 0.26998 | 0.7897 |
| Unknown177 | -1.4245 | 0.16832 |
| Unknown178 | 1.2767 | 0.21502 |
| Unknown179 | 1.735 | 0.096735 |
| Unknown18 | -1.6043 | 0.12292 |
| Unknown180 | -0.16026 | 0.87414 |
| Unknown181 | 0.16506 | 0.8704 |
| Unknown182 | 0.6512 | 0.52166 |
| Unknown183 | 0.0012328 | 0.99903 |
| Unknown184 | 0.018276 | 0.98558 |
| Unknown185 | -0.74904 | 0.46177 |
| Unknown186 | 1.0209 | 0.3184 |
| Unknown187 | 0.90877 | 0.37332 |
| Unknown188 | -0.25154 | 0.80373 |
| Unknown189 | 2.1939 | 0.039098 |
| Unknown19 | -1.6206 | 0.11935 |
| Unknown190 | -1.2658 | 0.21884 |
| Unknown191 | 0.68034 | 0.50338 |
| Unknown192 | 0.98873 | 0.33355 |
| Unknown193 | 0.36933 | 0.71541 |
| Unknown194 | -1.8453 | 0.078505 |
| Unknown195 | 0.75304 | 0.45941 |
| Unknown196 | -0.54979 | 0.588 |
| Unknown197 | -1.458 | 0.15897 |
| Unknown198 | 0.41831 | 0.67977 |
| Unknown199 | 0.46667 | 0.64532 |
| Unknown20 | -2.3935 | 0.025649 |
| Unknown200 | 2.022 | 0.055505 |
| Unknown201 | -0.0427 | 0.96633 |
| Unknown202 | 1.7712 | 0.090382 |
| Unknown203 | 0.52323 | 0.60604 |
| Unknown204 | 0.72349 | 0.477 |
| Unknown205 | 0.34519 | 0.73323 |
| Unknown206 | 0.084208 | 0.93365 |
| Unknown207 | 1.6359 | 0.11608 |
| Unknown208 | -1.1747 | 0.25268 |
| Unknown209 | 0.77685 | 0.44552 |
| Unknown21 | -0.47247 | 0.64124 |
| Unknown210 | 0.40399 | 0.69012 |
| Unknown211 | 0.30075 | 0.76643 |
| Unknown212 | 0.71735 | 0.48071 |
| Unknown213 | -0.55856 | 0.5821 |
| Unknown214 | 1.2048 | 0.24108 |
| Unknown215 | -0.22221 | 0.8262 |
| Unknown216 | -0.23652 | 0.81522 |
| Unknown217 | -0.90647 | 0.37451 |
| Unknown218 | 0.60478 | 0.55151 |
| Unknown219 | 0.24146 | 0.81143 |
| Unknown22 | -0.82946 | 0.41575 |
| Unknown220 | 1.0535 | 0.30353 |
| Unknown221 | -0.59217 | 0.55977 |
| Unknown222 | -0.90123 | 0.37723 |
| Unknown223 | -1.7495 | 0.094136 |
| Unknown224 | 0.33447 | 0.74119 |
| Unknown225 | 1.0447 | 0.30752 |
| Unknown226 | -1.5312 | 0.13998 |
| Unknown227 | -0.062354 | 0.95084 |
| Unknown228 | -0.11307 | 0.911 |
| Unknown229 | -1.2737 | 0.21606 |
| Unknown23 | -2.3326 | 0.029216 |
| Unknown230 | 0.72827 | 0.47413 |
| Unknown231 | 0.19821 | 0.8447 |
| Unknown232 | 1.0466 | 0.30667 |
| Unknown233 | -0.7384 | 0.46807 |
| Unknown234 | 0.35419 | 0.72657 |
| Unknown235 | -1.4349 | 0.16538 |
| Unknown236 | -2.0513 | 0.052334 |
| Unknown237 | 0.36737 | 0.71685 |
| Unknown238 | 0.77122 | 0.44878 |
| Unknown239 | 1.3648 | 0.18612 |
| Unknown24 | -1.2838 | 0.21258 |
| Unknown240 | 0.81054 | 0.42631 |
| Unknown241 | 1.2182 | 0.23606 |
| Unknown242 | -0.37137 | 0.71392 |
| Unknown243 | 1.1432 | 0.26525 |
| Unknown244 | -0.33318 | 0.74215 |
| Unknown245 | -1.3424 | 0.19317 |
| Unknown246 | 1.386 | 0.17962 |
| Unknown247 | 1.1791 | 0.25096 |
| Unknown248 | 0.30408 | 0.76392 |
| Unknown249 | 0.57577 | 0.57062 |
| Unknown25 | -1.5455 | 0.13649 |
| Unknown250 | -1.2702 | 0.21728 |
| Unknown251 | -0.49066 | 0.62853 |
| Unknown252 | 1.2521 | 0.22368 |
| Unknown253 | -0.15068 | 0.8816 |
| Unknown254 | 0.048818 | 0.96151 |
| Unknown255 | 0.10175 | 0.91987 |
| Unknown256 | 0.050166 | 0.96044 |
| Unknown257 | -0.84001 | 0.40994 |
| Unknown258 | 0.48051 | 0.63561 |
| Unknown259 | 0.69126 | 0.49664 |
| Unknown26 | -0.71626 | 0.48137 |
| Unknown260 | -0.19255 | 0.84907 |
| Unknown261 | 1.6932 | 0.10452 |
| Unknown262 | 1.3415 | 0.19344 |
| Unknown263 | 0.75037 | 0.46098 |
| Unknown264 | -2.3253 | 0.029672 |
| Unknown27 | -1.3264 | 0.19833 |
| Unknown28 | 0.001263 | 0.999 |
| Unknown29 | 0.71803 | 0.4803 |
| Unknown30 | 0.64689 | 0.5244 |
| Unknown31 | 0.47201 | 0.64157 |
| Unknown32 | -0.030875 | 0.97565 |
| Unknown33 | 0.66071 | 0.51566 |
| Unknown34 | -1.0087 | 0.32409 |
| Unknown35 | 0.98109 | 0.33722 |
| Unknown36 | -0.38948 | 0.70067 |
| Unknown37 | 0.86803 | 0.39475 |
| Unknown38 | 1.3598 | 0.18768 |
| Unknown39 | -0.20432 | 0.83998 |
| Unknown40 | 0.46523 | 0.64634 |
| Unknown41 | -0.12473 | 0.90187 |
| Unknown42 | 0.75123 | 0.46047 |
| Unknown43 | 1.3007 | 0.20682 |
| Unknown44 | 0.70804 | 0.48635 |
| Unknown45 | -0.9052 | 0.37516 |
| Unknown46 | 0.90495 | 0.3753 |
| Unknown47 | -0.97566 | 0.33984 |
| Unknown48 | 0.10278 | 0.91907 |
| Unknown49 | -0.15166 | 0.88084 |
| Unknown50 | 1.3789 | 0.18178 |
| Unknown51 | 1.8789 | 0.073576 |
| Unknown52 | 0.049398 | 0.96105 |
| Unknown53 | 0.86758 | 0.39499 |
| Unknown54 | 1.3608 | 0.18735 |
| Unknown55 | -1.0138 | 0.32172 |
| Unknown56 | 0.65991 | 0.51616 |
| Unknown57 | -0.28358 | 0.77938 |
| Unknown58 | -0.44889 | 0.6579 |
| Unknown59 | -0.12831 | 0.89907 |
| Unknown60 | 1.217 | 0.2365 |
| Unknown61 | 2.1562 | 0.042262 |
| Unknown62 | -0.74691 | 0.46302 |
| Unknown63 | -1.5984 | 0.12421 |
| Unknown64 | -0.16883 | 0.86747 |
| Unknown65 | -1.3321 | 0.19647 |
| Unknown66 | -0.74747 | 0.4627 |
| Unknown67 | 0.68049 | 0.50329 |
| Unknown68 | -0.0094582 | 0.99254 |
| Unknown69 | -1.0242 | 0.31686 |
| Unknown70 | -0.62644 | 0.53747 |
| Unknown71 | -1.0632 | 0.29924 |
| Unknown72 | 1.5359 | 0.13883 |
| Unknown73 | -0.074302 | 0.94144 |
| Unknown74 | 1.8019 | 0.085288 |
| Unknown75 | 1.5843 | 0.1274 |
| Unknown76 | 0.77248 | 0.44805 |
| Unknown77 | 0.64668 | 0.52453 |
| Unknown78 | 0.96614 | 0.34448 |
| Unknown79 | 2.4053 | 0.02501 |
| Unknown80 | 2.0509 | 0.052371 |
| Unknown81 | 1.1049 | 0.28114 |
| Unknown82 | 1.6836 | 0.10639 |
| Unknown83 | -0.79518 | 0.43501 |
| Unknown84 | 0.40287 | 0.69093 |
| Unknown85 | -0.095625 | 0.92468 |
| Unknown86 | 0.91523 | 0.36999 |
| Unknown87 | -1.1623 | 0.25757 |
| Unknown88 | -0.31327 | 0.75702 |
| Unknown89 | 0.80356 | 0.43025 |
| Unknown90 | 0.80304 | 0.43054 |
| Unknown91 | 0.1674 | 0.86859 |
| Unknown92 | 1.1439 | 0.26497 |
| Unknown93 | -0.7587 | 0.45608 |
| Unknown94 | 1.9815 | 0.060165 |
| Unknown95 | -1.4119 | 0.17198 |
| Unknown96 | -0.12794 | 0.89936 |
| Unknown97 | 0.088433 | 0.93033 |
| Unknown98 | 0.79155 | 0.43707 |
| Unknown99 | -1.4686 | 0.15609 |
